# Whole genome sequence association analysis of brain structural volume measures in the NHLBI TOPMed Program highlights novel loci in diverse participants

**DOI:** 10.1101/2025.06.11.25329426

**Authors:** Lincoln MP Shade, Mohsen Sharifitabar, Alexa Beiser, Claudia L Satizabal, Thomas H Mosley, Joanne E Curran, Jan Bressler, Susan R Heckbert, Timothy M Hughes, Thomas R. Austin, Ilya M Nasrallah, Lenore J Launer, Lisa R Yanek, Joshua C Bis, Harsha Doddapaneni, Richard A Gibbs, Stacey Gabriel, Namrata Gupta, Karine A Viaud-Martinez, Albert V Smith, Lauren A Opsasnick, Farrah Ammous, Jennifer A Smith, Donna K Arnett, Sharon L R Kardia, Bruce M Psaty, W T Longstreth, Rasika A Mathias, Paul Nyquist, Stephen S Rich, Jerome I Rotter, Eric Boerwinkle, Charles S DeCarli, David C Glahn, John Blangero, Myriam Fornage, David W Fardo, Sudha Seshadri, Chloé Sarnowski, the TOPMed Neurocognitive working group

## Abstract

Brain structural volumes are highly heritable and are linked to multiple neuropsychological outcomes, including Alzheimer’s disease (AD). Genome-wide association studies have successfully identified genetic variants associated with intracranial volume (ICV), total brain volume (TBV), hippocampal volume (HV), and lateral ventricular volume (LVV). However, these studies mostly focused on common genetic variants with minor allele frequencies (MAF) > 1%, and individuals included in most of these studies were of predominantly European ancestry. Here, we performed whole-genome sequence (WGS) association studies of MRI brain volumes in 7,674 individuals of diverse race and ethnicity from the Trans-Omics for Precision Medicine (TOPMed) program. We identified novel genetic loci on chromosomes 13 and 16 near *LINC00598* and *CACNG3* associated with HV and TBV, respectively (lead variants rs115674829, *P*-value = 1.7×10^-9^ in pooled analysis and rs150440001, *P*-value = 6.6×10^-9^ in black participants). Both lead variant minor A alleles are rarer in white participants (MAF = 0.14% and 0.03%) and in Hispanic participants (MAF = 1.5% and 0.17%) but more common in black participants (MAF = 13% and 1.5%). Rare variant aggregated analyses identified *RIPK1,* a gene encoding a kinase involved in neuroinflammation and promising target for AD treatment, suggestively associated with LVV (*P*-value=5×10^-6^). This study provides new insights into the genetic correlates of brain structural volumes and illustrates the importance of leveraging WGS data and cohorts of diverse race and ethnicity to better characterize the genetic architecture of complex polygenic traits.

## Background

Brain structural volumes are associated with a variety of important life and health outcomes, including educational attainment, schizophrenia, age of onset of Alzheimer’s disease, and cognitive ability.^1,2^ Intracranial volume (ICV) measures the volume of the cranium and represents a practical measure of the largest attainable total brain volume (TBV) during life. The hippocampi are bilateral structures in the medial cortex involved in memory formation, and decreased hippocampal volume (HV) is associated with both dementia and schizophrenia. The lateral ventricles are bilateral spaces within the cortex filled with cerebrospinal fluid. Lateral ventricular volume (LVV) increases throughout the course of aging, and LVV is enlarged in a variety of neurodegenerative diseases, including Alzheimer’s disease, Parkinson’s disease, and psychiatric diseases. Brain volumes are largely heritable, with heritability estimates ranging from 30 to 80% across different brain structural volumes, populations, and study methods.^3,4^ Therefore, characterizing the genetic architecture of brain volumes may help identify underlying contributors to Alzheimer’s disease and cognitive impairment.

Previous genome-wide association studies (GWAS) have identified genetic markers associated with ICV,^5–7^ HV,^8,9^ and LVV^10^ but have primarily focused on common genetic variants with minor allele frequencies (MAF) of greater than 1% and on participants of predominantly European ancestry. However, rare variants with MAF ≤1% may have outsized effects on human traits and disease risk.^11^ Furthermore, the focus on participants of predominantly European ancestry can cause studies to miss variants and genes associated with traits due to population differences in allele frequencies and linkage disequilibrium.^12^ Using phenotypes harmonized across studies with participants representing diverse race and ethnicity groups may improve the power to detect novel associations between less frequent genetic variations and brain volumes.

In this study, we sought to identify associations between both common (MAF>1%) and rare (MAF ≤1%) genetic variants and brain structural volumes (ICV, TBV, HV, and LVV) in participants of multiple race and ethnicity groups from eight studies as part of the NHLBI’s Trans Omics for Precision Medicine (TOPMed) Program.

## Methods

### Descriptions of studies

We included participants from eight TOPMed studies, listed in order of decreasing sample size: the Framingham Heart Study (FHS), the San Antonio Family Study (SAFS), the Atherosclerosis Risk in Communities study (ARIC), the Multi-Ethnic Study of Atherosclerosis (MESA), the Coronary Artery Risk Development in Young Adults Study (CARDIA), the Genetic Study of Atherosclerosis Risk (GeneSTAR), the Cardiovascular Health Study (CHS), and the Genetic Epidemiology Network of Arteriopathy (GENOA). Each of these studies have been described in detail previously, and brief descriptions of each study can be found in the Supplementary Note. Participants in these studies self-identified as at least one of four self-reported race or ethnicity groups (**Table 1**): white, African American or black, Hispanic, and Asian (specifically, Chinese) American. All studies underwent appropriate Institutional Review Board review, and all participants in each study provided informed consent.

**Table 1:** Summary and Demographics of TOPMed participants included in the WGS association analysis of brain volumes.

|  | ARIC | CARDIA | CHS | FHS | GeneSTAR | GENOA | MESA | SAFS | Overall |
| --- | --- | --- | --- | --- | --- | --- | --- | --- | --- |
| Sample Size | N=956 | N=786 | N=564 | N=2329 | N=580 | N=522 | N=917 | N=1020 | N=7674 |
| ICV, Mean (SD) | 1400 (157) | 1360 (152) | 1430 (154) | 1480 (142) | 1580 (154) | 1370 (130) | 1360 (145) | 1440 (148) | 1430 (160) |
| TBV, Mean (SD) | 1030 (108) | 1160 (125) | 965 (98.3) | 1130 (120) | 1260 (125) | 1040 (112) | 1090 (114) | 1050 (110) | 1100 (136) |
| HV, Mean (SD) | 6.97 (0.898) | 7.59 (0.831) | 6.92 (0.863) | 6.66 (0.745) | 9.95 (1.48) | NA (NA) | 7.11 (0.831) | 8.39 (0.919) | 7.36 (1.27) |
| LVV, Mean (SD) | 38.7 (20.1) | 18.0 (8.98) | 34.7 (16.8) | 22.7 (13.4) | 17.1 (9.01) | 20.5 (11.7) | 34.5 (18.4) | 13.1 (8.06) | 24.6 (16.5) |
| Age, Mean (SD) | 76.2 (5.27) | 51.3 (3.95) | 78.5 (4.07) | 59.3 (12.5) | 50.1 (10.7) | 61.5 (9.54) | 73.9 (8.05) | 38.7 (14.9) | 60.5 (16.2) |
| Female, N (%) | 563 (58.9%) | 419 (53.3%) | 324 (57.4%) | 1268 (54.4%) | 346 (59.7%) | 368 (70.5%) | 478 (52.1%) | 578 (56.7%) | 4344 (56.6%) |
| White, N (%) | 770 (80.5%) | 473 (60.2%) | 479 (84.9%) | 2329 (100%) | 316 (54.5%) | 0 (0%) | 373 (40.7%) | 8 (0.8%) | 4748 (61.9%) |
| Black, N (%) | 186 (19.5%) | 313 (39.8%) | 84 (14.9%) | 0 (0%) | 264 (45.5%) | 522 (100%) | 222 (24.2%) | 0 (0%) | 1591 (20.7%) |
| Hispanic, N (%) | 0 (0%) | 0 (0%) | 1 (0.2%) | 0 (0%) | 0 (0%) | 0 (0%) | 179 (19.5%) | 1009 (98.9%) | 1189 (15.5%) |
| Asian, N (%) | 0 (0%) | 0 (0%) | 0 (0%) | 0 (0%) | 0 (0%) | 0 (0%) | 143 (15.6%) | 3 (0.3%) | 146 (1.9%) |

FHS: the Framingham Heart Study, SAFS: the San Antonio Family Study, ARIC: the Atherosclerosis Risk in Communities study, MESA: the Multi-Ethnic Study of Atherosclerosis, CARDIA: the Coronary Artery Risk Development in Young Adults Study, GeneSTAR: the Genetic Study of Atherosclerosis Risk, CHS: the Cardiovascular Health Study, and GENOA: the Genetic Epidemiology Network of Arteriopathy.

### MRI protocol and phenotyping

MRIs were performed at individual study sites and underwent post-imaging processing by analysts at each study. Details on MRI procedures for each study are in **Supplementary Table 1**. To harmonize phenotypes between studies, we first converted all phenotype measurements to units of milliliters. Analysts from each study provided summaries of which volume segments were used to derive each phenotype (**Supplementary Table 2**). ICV and TBV were harmonized to include both supra- and infra-tentorial volumes for all studies, and ventricular volumes were excluded from TBV. HV and LVV reflected summed volumes for the left and right hippocampi and lateral ventricles, respectively. Details on our phenotype harmonization procedures can be found in the **Supplementary Note**.

### Whole genome sequencing (WGS)

WGS was performed as part of the TOPMed Program using DNA from whole blood collected in each study.^13^ Participants were sequenced at one of four sequencing centers and had an average coverage of 39X (see Supplementary Note for more details).^13^ We used TOPMed Freeze 8 that includes more than 80 different studies with approximately 158,000 samples and 1.02B genetic variants.

### Statistical analyses

In our primary analyses, we combined all available participants in a pooled analysis. In secondary analyses, we stratified participants by self-reported race and ethnicity and performed analyses separately in each race/ethnicity group with sufficient sample size (white, black, and Hispanic participants). We took this approach to compare results from the stratified analyses with the pooled analysis as black and Hispanic participants are often not included in genetic association studies of brain volumes. Stratified analyses were not done using Asian participants due to the relatively small available sample size. Secondary analyses also included additional adjustment for height for ICV analyses and conditional analyses on top signal in each region with genome-wide associations. LD calculations in the TOPMed data and regional association plots were done, within race/ethnicity group, using the Omics Analysis, Search and Information System (OASIS, https://omicsoasis.github.io).

WGS association analyses were performed on the NHLBI Biodata Catalyst Seven Bridges platform using the TOPMed Analysis Pipeline built using the GENESIS R package.^14^ For single-variant WGS association analyses, we used linear mixed-effects regression models (LMM) implemented in the SAIGE R package.^15^ Fixed-effect covariates included sex, age, age^2^, the top 10 genetic principal components (PCs), and study. Analyses of TBV, HV, and LVV were also adjusted for ICV. We also included a random effect for an empirical dense kinship matrix to account for relatedness between participants. Participants were excluded if they were noted to have dementia or large intracranial vascular disease at the time of MRI. We then ran null models using fixed effects covariates and removed participants whose residualized brain volumes were >3 SD away from the mean. Average Information Restricted Maximum Likelihood (AIREML) was used to fit LMM to account for potential heteroskedasticity between studies. We first regressed phenotypes on covariates, rank inverse normalized the residuals, and regressed on covariates again to create a null model for genetic association analysis. Score tests were then performed on individual genetic variants. *P*-values were calculated using a saddlepoint approximation.^15^ Genetic variants with a minor allele count (MAC) <20 were excluded from analysis. Genetic variants with *P*-values <1×10^-8^ were considered to have genome-wide significant associations, although we also report as “suggestive” variants that exceed the conventional genome-wide significance level for GWAS of common variants (*P*-value <5×10^-8^).

Previous GWAS have identified many significant associations between common genetic variants (MAF >1%) and brain structural volumes. We sought to confirm and potentially identify additional associations in recent brain volume GWAS loci, several of which have substantial participant overlap with the present study, and to evaluate if the associations differ across race/ethnicity groups. We used large available GWAS for ICV, HV, and LVV.^5–10,16^ Most of these studies used a single-variant genome-wide significance level of *P*-value < 5×10^-8^ and reported lead genetic variants at each independent locus significantly associated with the brain volume trait studies. In one study of 44 hippocampal subvolumes, a threshold of *P*-value=1.14×10^-9^ (5×10^-8^ / 44) was used.^16^ For this study, we used only variants found to be associated with either whole left or right hippocampal volumes, as the genetic factors of hippocampal subvolumes differ from whole hippocampal volumes.^17^ We compared the results of lead variants in these studies to results from our pooled analyses. We considered variant association to be replicated in our pooled single-variant analysis if the effect direction was concordant to the previously reported association and if the variant had a *P*-value <0.05. We also examined the results of the same lead variants in our race/ethnicity-stratified analyses to provide additional context on previously reported associations across race and ethnicity groups.

To improve power for detecting associations between rare functional variants and brain MRI volumes, we then performed aggregate association tests in GENESIS using the optimal sequence-kernel association test (SKAT-O), which performs a weighted combination of the SKAT and burden tests.^18^ Only variants with MAF ≤1% in our sample and that had Variant Effect Predictor (VEP) high-confidence loss-of-function (LOF) or missense annotations were used.^19^ SKAT-O ⍴ values (weights) tested included [0, 1] in steps of 0.1. Genes with fewer than 10 total minor alleles present across rare LOF and missense variants in our study participants were excluded from analysis. Gene (aggregation unit) associations with Bonferroni-corrected *P*-values < 2.5×10^-6^ (corresponding to 0.05/20,000 genes) were considered to be significant, and gene associations with *P*-values < 5×10^-5^ were considered to be suggestive.

To identify loci exhibiting potential functional molecular biological effect, we performed Bayesian colocalization analysis using quantitative trait loci (QTL) data from three resources: expression (eQTL) and splicing (sQTL) QTL in 48 tissues from the Genotype Tissue Expression Project (GTEx) v8 release,^20^ eQTL and methylation (mQTL) in the brain dorsolateral prefrontal cortex from the Religious Orders Study and Rush Memory and Aging Project (ROSMAP) xQTL study,^21^ and eQTL in six tissues (whole blood, lung, nasal epithelium, T-cells, monocytes, and peripheral blood mononuclear cells [PBMC]) from TOPMed.^22^ All variants in the pooled single-variant WGS association analyses with *P*-values <10^-6^ were first checked for QTL status in each study. Colocalization analyses were then performed using the coloc R package with default prior probabilities.^23^ A posterior probability of colocalization (PrC) > 80% was considered evidence of colocalization.

## Results

We analyzed up to 7,674 participants (white N = 4,748; Hispanic N = 1,189; black N = 1,591; Asian N=146) from eight different TOPMed studies (**Table 1**). Self-reported race and ethnicity clustered by genetic PC1 and PC2 (**Supplementary Figure 1**). The majority of our participants were female (56.6%). Mean age at MRI varied between studies, from 38.7 years in SAFS to 78.5 years in CHS, with the pooled mean age at MRI for participants being 60.5 (SD 16.2) years.

### Single-variant associations

Manhattan plots for the pooled analysis for each brain structural volume are presented in **Figure 1**. Manhattan plots for the race/ethnicity stratified analyses and QQ plots for the pooled analyses are presented in **Supplementary Figures 2-5**. Genomic inflation lambda values for studied traits ranged from 1.013-1.025, indicating controlled type I error and low risk of bias due to population stratification. Main association signals are presented in **Table 2**. One novel locus on Chromosome 13 was significantly associated with HV in the pooled analysis (lead variant rs115674829, beta = -0.27 mL, *P*-value = 1.7×10^-9^, MAF = 0.02). When we stratified by race/ethnicity, we detected one locus on Chromosome 16 associated with TBV at genome-wide significance in black participants (rs150440001, beta = 37.13, *P*-value = 6.59×10^-9^, MAF = 1.5%). Two previously reported loci on Chromosome 6 were significantly associated with ICV (**Supplementary Figure 6**). The top associated SNPs were rs35947181 (beta = -14 mL, *P*-value = 4.9×10^-12^, MAF = 0.47) and rs1490384 (beta =12 mL, *P*-value = 2.6×10^-9^, MAF = 0.49).

**Figure 1:**
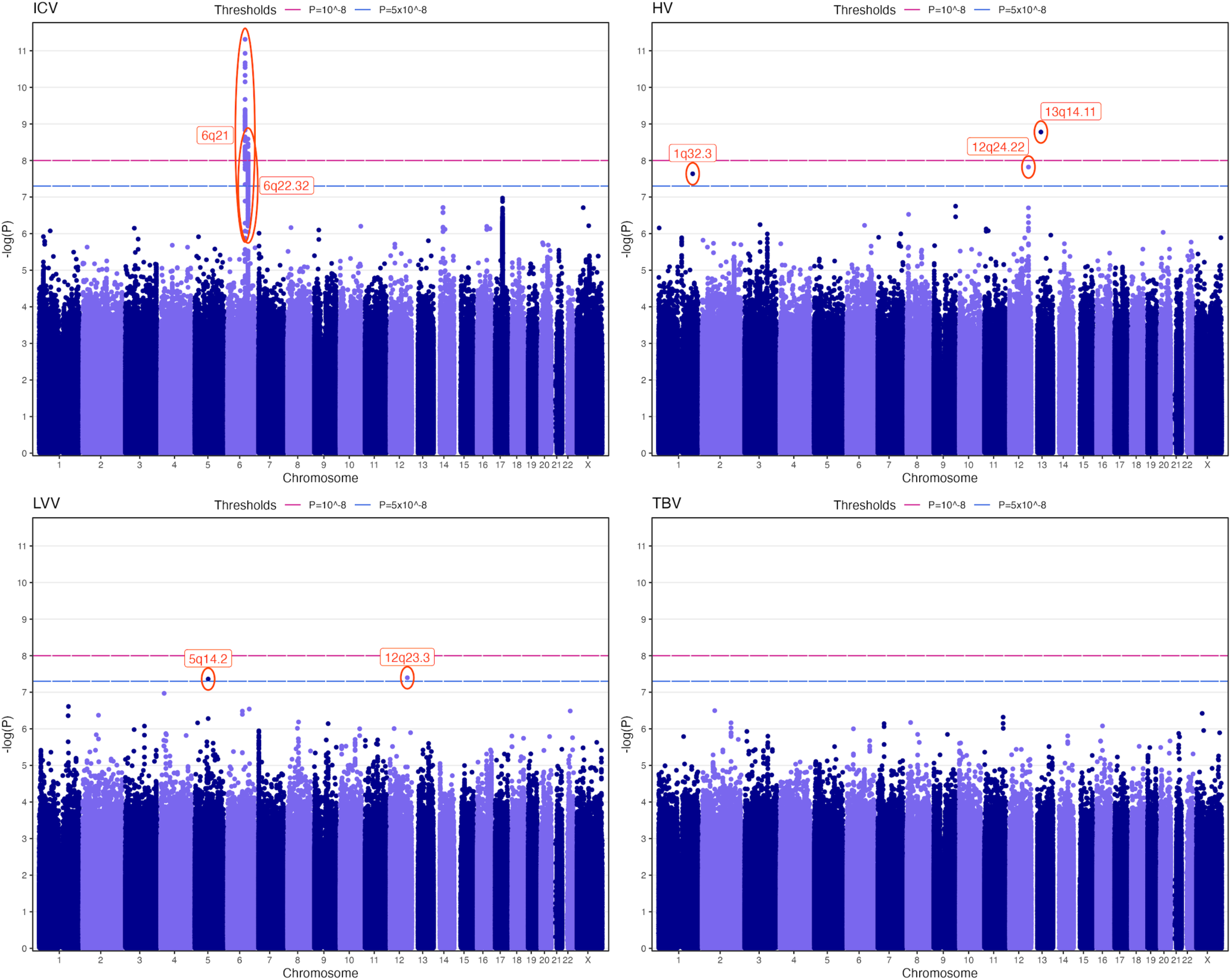
Manhattan Plots for Brain MRI volumes associations based on TOPMed Whole-Genome Sequence association analysis. Blue dashed line demarcates sub-genome-wide significance (P ≤ 5×10^-8^). Pink dashed line demarcates genome-wide significance (P ≤ 10^-8^). HV = hippocampal volume; ICV = intracranial volume; LVV = log lateral ventricular volume; TBV = total brain volume. Only genetic variants passing minor allele count (MAC) ≥ 20 are represented.

**Table 2:** Main genetic variants associated with brain volumes in single-variant association analyses using TOPMed whole genome sequence data.

|  |  |  |  |  |  | Pooled analysis (N=7,040-7,674) |  |  |  | Black analysis (N=1,015-1,591) |  |  |  | White analysis (N=4,682-4,740) |  |  |  | Hispanic analysis (N=1,188) |  |  |  |
| --- | --- | --- | --- | --- | --- | --- | --- | --- | --- | --- | --- | --- | --- | --- | --- | --- | --- | --- | --- | --- | --- |
| Trait | Chr | Pos b38 (Mb) | rsid | ref/alt | Gene | FREQ | BETA | SE | P | FREQ | BETA | SE | P | FREQ | BETA | SE | P | FREQ | BETA | SE | P |
| TBV | 16 | 24.4 | rs150440001 | G/A | *CACNG3* | 0.004 | 26.11 | 5.78 | 6.3E-06 | 0.015 | 37.13 | 6.40 | 6.6E-09 | 0.0003 | NA | NA | NA | 0.002 | NA | NA | NA |
| LVV | 12 | 106.1 | rs12146713 | T/C | *NUAK1* | 0.067 | 0.07 | 0.01 | 4.0E-08 | 0.016 | 0.02 | 0.06 | 0.77 | 0.093 | 0.08 | 0.01 | 1.2E-07 | 0.041 | 0.06 | 0.04 | 0.16 |
| LVV | 5 | 82.7 | rs78861128 | C/T | *LOC124901020* | 0.273 | 0.04 | 0.01 | 4.4E-08 | 0.219 | 0.07 | 0.02 | 2.0E-05 | 0.280 | 0.03 | 0.01 | 4.0E-04 | 0.313 | 0.04 | 0.02 | 0.06 |
| HV | 1 | 211.5 | rs567130812 | G/C | *RD3* | 0.003 | 0.61 | 0.11 | 2.3E-08 | 0.000 | NA | NA | NA | 0.004 | 0.61 | 0.11 | 1.7E-08 | 0.0004 | NA | NA | NA |
| HV | 12 | 104.5 | rs17811260* | T/C | *CHST11* | 0.024 | 0.05 | 0.04 | 0.16 | 0.029 | 0.55 | 0.10 | 1.7E-7 | 0.024 | -0.04 | 0.04 | 0.41 | 0.019 | 0.05 | 0.10 | 0.61 |
| HV | 12 | 107.0 | rs1053051 | T/C | *TMEM263* | 0.588 | 0.003 | 0.012 | 0.81 | 0.779 | -0.24 | 0.04 | 2.7E-08 | 0.503 | 0.03 | 0.01 | 0.017 | 0.757 | 0.01 | 0.03 | 0.72 |
| HV | 12 | 116.8 | rs4767458 | T/C | *RNFT2* | 0.922 | 0.10 | 0.02 | 7.0E-06 | 0.932 | 0.17 | 0.07 | 0.02 | 0.912 | 0.10 | 0.02 | 1.8E-05 | 0.947 | 0.04 | 0.06 | 0.50 |
| HV | 12 | 116.9 | rs146607495 | C/T | *HRK* | 0.082 | 0.12 | 0.02 | 1.5E-08 | 0.023 | 0.15 | 0.12 | 0.22 | 0.104 | 0.10 | 0.02 | 8.4E-06 | 0.058 | 0.21 | 0.06 | 3.0E-04 |
| HV | 13 | 40.3 | rs115674829 | G/A | *LINC00598* | 0.022 | -0.27 | 0.04 | 1.7E-09 | 0.128 | -0.27 | 0.05 | 4.9E-07 | 0.0014 | NA | NA | NA | 0.015 | -0.34 | 0.11 | 0.003 |
| ICV | 18 | 44.4 | rs148196235 | AC/A | *LINC01478* | 0.110 | 4.26 | 2.88 | 0.14 | 0.090 | 37.70 | 6.76 | 2.4E-08 | 0.115 | -3.64 | 3.56 | 0.306 | 0.123 | 7.91 | 7.46 | 0.29 |
| ICV | 20 | 45.4 | rs6032173 | A/C | *PIGT* | 0.203 | 10.63 | 2.24 | 2.0E-06 | 0.185 | 3.86 | 4.97 | 0.44 | 0.198 | 15.87 | 2.85 | 2.6E-08 | 0.218 | -2.76 | 5.74 | 0.63 |
| ICV | 6 | 108.5 | rs9480861 | C/T | *LOC124901370* | 0.465 | -10.05 | 1.90 | 1.3E-07 | 0.725 | -14.24 | 4.36 | 1.0E-03 | 0.434 | -9.09 | 2.31 | 8.6E-05 | 0.275 | -6.75 | 5.48 | 0.22 |
| ICV | 6 | 108.6 | rs768023 | G/A | *LOC124901370* | 0.512 | 11.83 | 1.91 | 6.4E-10 | 0.191 | 11.72 | 5.18 | 0.024 | 0.603 | 12.68 | 2.30 | 3.7E-08 | 0.559 | 9.69 | 4.87 | 0.047 |
| ICV | 6 | 108.7 | rs35947181 | G/GC | *FOXO3* | 0.474 | -13.61 | 1.97 | 4.9E-12 | 0.840 | -19.00 | 5.57 | 6.4E-04 | 0.365 | -13.34 | 2.38 | 2.0E-08 | 0.439 | -14.91 | 4.85 | 0.002 |
| ICV | 6 | 126.5 | rs1490384 | C/T | *LOC105377989* | 0.490 | 11.63 | 1.95 | 2.6E-09 | 0.204 | 14.66 | 4.96 | 3.0E-03 | 0.519 | 11.67 | 2.30 | 4.0E-07 | 0.701 | 6.72 | 5.45 | 0.218 |

HV = hippocampal volume; ICV = intracranial volume; LVV = log lateral ventricular volume; TBV = total brain volume.
\*secondary association identified by conditional analysis (BETA=0.57, P=4.2E-8 after conditioning on variant rs1053051 in black analysis)

Two additional loci were associated with HV in the pooled analysis at a near genome-wide level: a novel intronic locus of *RD3* on Chromosome 1 (rs567130812, beta = 0.61 mL, *P*-value = 2.3×10^-8^, MAF = 0.28%) and a previously identified non-coding locus of *HRK* on Chromosome 12 (rs146607495, beta = 0.12 mL, *P*-value = 1.5×10^-8^, MAF = 8.2%; **Supplementary Figure 7**). Interestingly, the lead variant in *HRK* has a low frequency in black participants (MAF=2.3%) and no association is observed in this group (*P*-value = 0.22). Two loci were associated with LVV at a near genome-wide level: a novel locus on Chromosome 5 (rs78861128, beta = 0.04 log-mL, *P*-value = 4.3×10^-8,^ MAF = 27%; **Supplementary Figure 8**), and a previously reported locus on Chromosome 12 (rs12146713, beta = 0.07 log-mL, *P*-value = 4.0×10^-8,^ MAF = 6.7%; **Supplementary Figure 9**). The lead variant at 12q23 has a lower frequency in black and Hispanic participants (1.6% and 4.1% respectively) and no association is observed in these groups (*P*-values = 0.77 and 0.16 respectively). No variants were significantly associated with TBV in the pooled analysis.

Several loci in the race/ethnicity-stratified analyses reached a near-genome-wide level of association with brain regional volumes. A novel locus on Chromosome 12 in the 3’ untranslated region of *TMEM263* was associated with HV only in black participants (rs1053051, beta = 0.24 mL, *P*-value = 2.7×10^-8^, MAF = 22%; **Supplementary Figure 10**). A secondary association signal was identified in *CHST11* by conditional analysis (rs17811260, beta=0.57 mL, *P*-value = 4.2×10^-8^, MAF = 2.9%; **Supplementary Figure 11**). A novel locus on Chromosome 18 in the long intergenic non-coding RNA gene *LINC01478* was associated with ICV only in black participants (rs148196235, beta = 37.71, *P*-value = 2.38×10^-8^, MAF = 9%; **Supplementary Figure 12**).

In the analysis of white participants, the same *RD3* locus identified in the pooled analysis was associated with HV (rs567130812, beta = 0.61 mL, *P*-value = 1.7×10^-8^, MAF = 0.40%). This locus was almost exclusively found in white participants, with MAC of one in Hispanic and MAC of zero in black participants. A novel locus on Chromosome 20 in *PIGT* gene was associated with ICV only in white participants (rs6032173, beta = 15.87, *P*-value = 2.63×10^-8^, MAF = 20%; **Supplementary Figure 10**). No novel associations were detected in the Hispanic stratified analyses.

Among 71 previously reported brain volume loci, we replicated 20 previously identified HV-associated loci, five ICV-associated loci, and seven LVV-associated loci at a nominal level of statistical significance (p <0.05, **Supplementary Table 3**).

### Gene-based aggregate analyses of rare-variants

No genes were significantly associated at *P*-value < 2.5×10^-6^ with any of the brain MRI volumes in aggregate association tests using LOF and missense variants (**Supplementary Tables 4-7**). However, we detected two genes at a suggestive level of *P*-value<5×10^-5^: *RIPK1* associated with LVV (P_min_=5×10^-6^)^24–26^ and *BIRC6* associated with ICV (P_min_=1.8×10^-5^).

### Colocalization analyses with QTL studies

Two HV-associated loci showed evidence of colocalization with GTEx eQTLs. The Chromosome 12 *HRK* locus with lead variant rs146607495 (*P*-value = 1.5×10^-8^) colocalized with *FBXO21* in the left ventricle of the heart (PrC = 83%; **Supplementary Figure 13**), and a suggestive locus on Chromosome 8 with lead variant rs12680137 (*P*-value = 3.0×10^-7^) colocalized with expression of ENSG00000280273, a pseudogene that transcribes RNA that is antisense to *MTMR9*, in the caudate nucleus of the basal ganglia (PrC = 85%), **Supplementary Figure 14.** A locus suggestively associated with LVV on Chromosome 1 with lead variant rs3795503 (*P*-value = 2.4×10^-7^) showed evidence of colocalization with *KIAA1614* expression across 20 tissues, including the brain frontal cortex in both GTEx (PrC = 93%) and ROSMAP (PrC >99%; **Supplementary Figure 15**).

## Discussion

We studied the whole genome-wide contributors to MRI-measured brain structural volumes using 7,674 TOPMed participants of diverse race and ethnicity. We identified a novel locus on Chromosome 13q14 associated with HV in the pooled sample. In stratified analyses 16p12 associated with TBV in black participants and a novel locus on Chromosome 20 in *PIGT* gene was associated with ICV only in white participants. Gene-based rare variant aggregation tests did not reveal any significant associations, although two suggestive associations (*RIPK1* and *BIRC6* genes) in the pooled sample merit further consideration. Finally, we confirmed 32 genetic loci previously associated with brain MRI volume measurements at a significance level of *P*-value <0.05. These replications include 20 HV-associated loci found in a multi-ancestry study (European and East Asian) that did not include sample overlap with the present study.^16^

In the pooled sample, Chromosome 13q14 associated with HV. The lead variant at this locus, rs115674829, is located within the long intergenic non-coding RNA gene *LINC00598*, which has been identified to have associations with multiple traits in prior GWAS, with its strongest associations being with monocyte count and body height. In the RegulomeDB resource,^27,28^ the rs115674829 effect allele is associated with chromatin accessibility levels in the head of the caudate nucleus and the posterior cingulate gyrus.

The genetic variant rs150440001 (16p12) associated with TBV in black participants. The lead variant is located in the intron 2 of *CACNG3 (calcium voltage-gated channel auxiliary subunit gamma)*. *CACNG3* was among the top six genes, all related to “ion transport”, for which a negative relationship of protein expression level with AD neuropathologic changes was reported.^29^ These results suggest that dysregulation of ion transport across brain regions might play a critical role in AD pathology.^29^

A novel locus on Chromosome 20 in *PIGT* gene was associated with ICV only in white participants (rs6032173, *P*-value = 2.63×10^-8^, MAF = 20%). The lead genetic variant is common (MAF > 18%) in all population groups. This locus has not been reported by previous GWAS of ICV and was not present in the summary statistics of these studies,^5,6^ as it was not present in the imputation panels (1000 Genomes reference panel phase 1 version 3, Haplotype Reference Consortium and the UK10K) or did not pass imputation quality filters. The regional association plot (**Supplementary Figure 10**) shows that no SNPs are in strong LD (r2>0.6) with the lead variant which would make the imputation difficult if the variant is not present on the genotyping chip. This result highlights the importance of leveraging WGS data. The lead variant rs6032173 is an expression QTL (associated with a decreased expression) in blood for two genes in the region, *SYS1* and *TP53TG5*.^20,30^

Our gene-based analyses of rare variants identified the receptor interacting serine/threonine kinase 1 (*RIPK1*) suggestively associated with LVV. This gene encodes a protein that plays a role in inflammation and cell death in response to tissue damage. RIPK1/RIPK3 kinase-mediated necrosis is referred to as necroptosis. RIPK1 plays a crucial role in maintaining cellular and tissue homeostasis by integrating inflammatory responses and cell death signaling pathways including apoptosis and necroptosis, which have been implicated in diverse physiological and pathological processes. Suppression of RIPK1 activation can help inhibit multiple cell death pathways, ameliorate neuroinflammation, and restrain pathological progression of many human diseases. Necroptosis promotes further cell death and neuroinflammation in the pathogenesis of several neurodegenerative diseases, including multiple sclerosis, amyotrophic lateral sclerosis, Parkinson disease and AD.^24,26^ Inhibiting RIPK1 kinase has been shown to mitigate brain injury after subarachnoid hemorrhage through attenuation of RIPK1-driven neuroinflammation and neuronal apoptosis in both in vivo and in vitro models.^25^ Investigation on RIPK1 as a promising target for AD treatment is currently ongoing.^26^

Our gene-based analyses of rare variants also identified baculoviral IAP repeat containing 6 (*BIRC6*) suggestively associated with ICV. This gene encodes a protein that inhibits apoptosis by facilitating the degradation of apoptotic proteins by ubiquitination.^31,32^ While most research on *BIRC6* relates to cancer, inhibition of *BIRC6* has been shown to repress autophagosome-lysosome fusion in neuronal axons leading to axonal dystrophy in Alzheimer’s disease mouse models.^33^

One variant suggestively associated with LVV in the present study, rs3795503, is a missense variant (N>K) for KIAA1614. *KIAA1614* codes for the uncharacterized protein KIAA1614. While literature on *KIAA1614* is scarce, it has been associated with multiple other brain phenotypes including fractional anisotropy and cortical surface area.^34,35^ rs3795503 is a strong eQTL for *KIAA1614* in the brain cortex in both GTEx and ROSMAP, and is also an mQTL in ROSMAP. Our results indicate that additional research on the relationship between *KIAA1614* and brain structures is merited.

For two of our novel loci (13q14 and 16p12), we note that although the lead variants were not rare in the pooled sample, the MAFs showed substantial differentiation between race/ethnicity groups. For both, the minor alleles were more common in black (MAF = 13% for rs115674829 and MAF = 2% for rs150440001) vs Hispanic (MAF = 1.5% and 0.2%, respectively) and white (MAF = 0.14% and 0.03%, respectively) participants. These loci were thus likely not identified in previous GWAS of HV and TBV because those studies solely included participants of groups with predominantly European ancestry and studied genetic variants with MAF ≥1%,^8,9^ meaning that these variants were not studied/included due to the rare MAF or low MAC in those participants.

Particular strengths of this study include the use of WGS in lieu of genotyping arrays to enable the study of intermediate frequency and rare genetic variants and the inclusion of participants of multiple races and ethnicities. While race and ethnicity may not accurately reflect underlying genetic ancestry, we found stratification by these variables appropriate for the current study because participants self-identified with these groups and broadly clustered by these groups when plotting genetic PC1 and PC1 (see **Supplementary Figure 1**). Our study represents the largest study of brain volumes in black participants (∼180% increase in sample size compared to Hibar *et al* (2017) GWAS of HV),^9^ and the first to our knowledge to include black and Hispanic participants in a discovery rather than a replication cohort.

Our study has several limitations. Our available sample sizes, ranging from 7,040 participants for HV to 7,674 for ICV, were smaller than the sample sizes used in recent GWAS of brain MRI volumes. And while our study includes more racial and ethnic diversity than prior studies, the available sample sizes for Hispanic and black participants in our study were not as well-powered for identifying loci associated with brain MRI volumes as the white sample. We also note that there is a substantial overlap between our study and previous GWAS of brain MRI volumes. In particular, FHS, ARIC, GeneSTAR, and CHS are part of the Cohorts for Heart and Aging Research in Genetic Epidemiology (CHARGE) consortium, which has led many of the previous GWAS of brain MRI volumes. Therefore, our results do not constitute independent replications of previous results.

In conclusion, we leveraged eight studies of diverse TOPMed participants to study MRI-derived brain structural volumes using whole-genome sequencing. We identify multiple novel genetic loci associated with brain volumes and replicate over thirty known loci from prior studies. Our results demonstrate the importance of leveraging whole-genome sequence data and genetically diverse populations for better characterizing the genetic architecture of complex traits such as brain volumes.

## Availability of data and materials

Data for each participating study can be accessed through the database of Genotypes and Phenotypes (dbGaP) with the corresponding accession numbers (ARIC, phs001211; CARDIA, phs001612; CHS, phs001368; FHS, phs000974; GeneSTAR, phs001218; GENOA, phs001345; MESA, phs001416; SAFS, phs001215).

## Funding

### TOPMed

Molecular data for the Trans-Omics in Precision Medicine (TOPMed) program was supported by the National Heart, Lung and Blood Institute (NHLBI). Study specific omics support information is provided in the **Supplement**. Core support including centralized genomic read mapping and genotype calling, along with variant quality metrics and filtering were provided by the TOPMed Informatics Research Center (3R01HL-117626-02S1; contract HHSN268201800002I). Core support including phenotype harmonization, data management, sample-identity QC, and general program coordination were provided by the TOPMed Data Coordinating Center (R01HL-120393; U01HL-120393; contract HHSN268201800001I). Study-specific funding information is included in the Supplement. The Analysis Commons was funded by R01HL131136, R01HL105756.

### Individual

L.M.P.S. was funded by T32 AG57461 and National Institute of Neurological Disorders and Stroke (NINDS) F30 NS124136. M.F. was funded by U01AG058589. A.B. was funded by R01 AG054076 and R01 AG016495. T.M.H. is funded by grants U01 HL096812, R01 AG054069, and R01 AG058969 from the National Institute on Aging (NIA). C.L.S. was funded by R01 AG059727, R01 AG082360, UO1 NS125513, and P30 AG066546-6181. D.W.F. was funded by RF1 AG082339. C.S. was funded by R00 AG066849. S.S. was funded by R01 HL105756, R01 AG033193, P30 AG066546, RF1 AG059421, R01 AG054076, and R01 AG049607. C.S.D. received support from U19 NS120384, P30 AG07297, R01 AG075758, RF1 AG077639 and RF1 NS130659. M.S was supported by P30 AG066546, R01 AG082360, and R21 AG087907.

## Acknowledgements

We gratefully acknowledge the studies and participants who provided biological samples and data for TOPMed. We acknowledge the TOPMed Program. Study-specific acknowledgements are included in the Supplement.

## References

1. Braskie, M. N., Ringman, J. M. & Thompson, P. M. Neuroimaging measures as endophenotypes in Alzheimer’s disease. Int. J. Alzheimers Dis. 2011, 490140 (2011).

2. Lee, J. J., McGue, M., Iacono, W. G., Michael, A. M. & Chabris, C. F. The causal influence of brain size on human intelligence: Evidence from within-family phenotypic associations and GWAS modeling. Intelligence 75, 48–58 (2019).

3. Jansen, A. G., Mous, S. E., White, T., Posthuma, D. & Polderman, T. J. C. What Twin Studies Tell Us About the Heritability of Brain Development, Morphology, and Function: A Review. Neuropsychol. Rev. 25, 27–46 (2015).

4. Zhao, B. et al. Heritability of Regional Brain Volumes in Large-Scale Neuroimaging and Genetic Studies. Cereb. Cortex N. Y. NY 29, 2904–2914 (2019).

5. Ikram, M. A. et al. Common variants at 6q22 and 17q21 are associated with intracranial volume. Nat. Genet. 44, 539–544 (2012).

6. Adams, H. H. H. et al. Novel genetic loci underlying human intracranial volume identified through genome-wide association. Nat. Neurosci. 19, 1569–1582 (2016).

7. Nawaz, M. S. et al. Thirty novel sequence variants impacting human intracranial volume. Brain Commun. 4, fcac271 (2022).

8. Bis, J. C. et al. Common variants at 12q14 and 12q24 are associated with hippocampal volume. Nat. Genet. 44, 545–551 (2012).

9. Hibar, D. P. et al. Novel genetic loci associated with hippocampal volume. Nat. Commun. 8, 13624 (2017).

10. Vojinovic, D. et al. Genome-wide association study of 23,500 individuals identifies 7 loci associated with brain ventricular volume. Nat. Commun. 9, 3945 (2018).

11. Cirulli, E. T. & Goldstein, D. B. Uncovering the roles of rare variants in common disease through whole-genome sequencing. Nat. Rev. Genet. 11, 415–425 (2010).

12. Wojcik, G. L. et al. Genetic analyses of diverse populations improves discovery for complex traits. Nature 570, 514–518 (2019).

13. Taliun, D. et al. Sequencing of 53,831 diverse genomes from the NHLBI TOPMed Program. Nature 590, 290–299 (2021).

14. Conomos, M. P., et al. GENESIS: GENetic EStimation and Inference in Structured Samples (GENESIS): Statistical Methods for Analyzing Genetic Data from Samples with Population Structure and/or Relatedness. (2019).

15. Zhou, W. et al. Efficiently controlling for case-control imbalance and sample relatedness in large-scale genetic association studies. Nat. Genet. 50, 1335–1341 (2018).

16. Liu, N. et al. Cross-ancestry genome-wide association meta-analyses of hippocampal and subfield volumes. Nat. Genet. 55, 1126–1137 (2023).

17. van der Meer, D. et al. Brain scans from 21,297 individuals reveal the genetic architecture of hippocampal subfield volumes. Mol. Psychiatry 25, 3053–3065 (2020).

18. Lee, S., Wu, M. C. & Lin, X. Optimal tests for rare variant effects in sequencing association studies. Biostat. Oxf. Engl. 13, 762–775 (2012).

19. McLaren, W. et al. The Ensembl Variant Effect Predictor. Genome Biol. 17, 122 (2016).

20. GTEx Consortium et al. Genetic effects on gene expression across human tissues. Nature 550, 204–213 (2017).

21. Ng, B. et al. An xQTL map integrates the genetic architecture of the human brain’s transcriptome and epigenome. Nat. Neurosci. 20, 1418–1426 (2017).

22. Liu, C. et al. Whole Genome DNA and RNA Sequencing of Whole Blood Elucidates the Genetic Architecture of Gene Expression Underlying a Wide Range of Diseases. 2022.04.13.22273841 Preprint at 10.1101/2022.04.13.22273841 (2022).

23. Giambartolomei, C. et al. Bayesian Test for Colocalisation between Pairs of Genetic Association Studies Using Summary Statistics. PLOS Genet. 10, e1004383 (2014).

24. Yuan, J., Amin, P. & Ofengeim, D. Necroptosis and RIPK1-mediated neuroinflammation in CNS diseases. Nat. Rev. Neurosci. 20, 19–33 (2019).

25. Wu, Y. et al. Inhibiting RIPK1-driven neuroinflammation and neuronal apoptosis mitigates brain injury following experimental subarachnoid hemorrhage. Exp. Neurol. 374, 114705 (2024).

26. Li, S., Qu, L., Wang, X. & Kong, L. Novel insights into RIPK1 as a promising target for future Alzheimer’s disease treatment. Pharmacol. Ther. 231, 107979 (2022).

27. Boyle, A. P. et al. Annotation of functional variation in personal genomes using RegulomeDB. Genome Res. 22, 1790–1797 (2012).

28. Dong, S. et al. Annotating and prioritizing human non-coding variants with RegulomeDB. 2022.10.18.512627 Preprint at 10.1101/2022.10.18.512627 (2022).

29. Jia, Y. et al. Proteomic and Transcriptomic Analyses Reveal Pathological Changes in the Entorhinal Cortex Region that Correlate Well with Dysregulation of Ion Transport in Patients with Alzheimer’s Disease. Mol. Neurobiol. 58, 4007–4027 (2021).

30. Võsa, U. et al. Large-scale cis- and trans-eQTL analyses identify thousands of genetic loci and polygenic scores that regulate blood gene expression. Nat. Genet. 53, 1300–1310 (2021).

31. Liu, S.-S. et al. Molecular mechanisms underlying the BIRC6-mediated regulation of apoptosis and autophagy. Nat. Commun. 15, 891 (2024).

32. Saleem, M. et al. Inhibitors of Apoptotic Proteins: New Targets for Anticancer Therapy. Chem. Biol. Drug Des. 82, 243–251 (2013).

33. Zhang, L. et al. BRUCE silencing leads to axonal dystrophy by repressing autophagosome-lysosome fusion in Alzheimer’s disease. Transl. Psychiatry 11, 421 (2021).

34. Fan, C. C. et al. Multivariate genome-wide association study on tissue-sensitive diffusion metrics highlights pathways that shape the human brain. Nat. Commun. 13, 2423 (2022).

35. Sha, Z., Schijven, D., Fisher, S. E. & Francks, C. Genetic architecture of the white matter connectome of the human brain. Sci. Adv. 9, eadd2870 (2023).

