## Supplementary Note for "Whole genome sequence association analysis of brain structural volume measures in the NHLBI TOPMed Program highlights novel loci in diverse participants"

**Supplementary Information**

### TOPMed WGS data

Studies have been added to the TOPMed program in approximately five yearly “Phases”, with Phase 1 beginning in October 2014, and Phase 5 in October 2018. TOPMed WGS data are acquired and reads aligned to the reference genome continuously by multiple sequencing centers. The TOPMed Informatics Research Center (IRC) performs periodically variant identification and genotype calling on all samples available at a given time and the resulting call set is referred to as a genotype “Freeze”.
Briefly, approximately 30X WGS was performed at several different sequencing centers using DNA from blood and Illumina HiSeq X technology. In most cases, all samples for a given study within a given phase were sequenced at the same center. The reads were aligned to human genome build GRCh38 using a common pipeline across all centers. The IRC performed joint genotype calling on all samples in Freeze 8. The resulting VCF files were split by study and consent group for distribution to approved dbGaP users. They can be reassembled easily for cross-study, pooled analysis since the files for all studies contain identical lists of variant sites. Quality control was performed at each stage of the process by the sequencing centers, the IRC and the TOPMed Data Coordinating Center (DCC). Only samples that passed QC were included in the call set, whereas all variants (whether passed or failed) were included.

### Brain Volumes Phenotype Harmonization

TBV from two studies did not include the same volumes as the other studies: TBV in GeneSTAR included ventricular volumes but did not include infratentorial volume, and TBV in GENOA included ventricular volumes. To attempt to improve harmonization between studies, we created two imputation models. First, we created an imputation model of TBV from FHS (r2=0.989), which had ventricular volumes and supra-tentorial TBV available, in order to impute infratentorial volume in GenSTAR. This model amounted to linearly scaling GeneSTAR TBV and adding a fixed volume, and the resulting volume was highly correlated with supratentorial TBV (r2 >0.999). We then created two separate models to impute ventricular volume-free TBV using FHS and SAFS, which had both measures available. Both models successfully imputed ventricular volume-free TBV (r2 >0.999 between actual and imputed phenotypes) and the coefficients in each model were similar, so we used the model based on SAFS to impute TBV in GeneSTAR and GENOA.

### Study Descriptions

#### ARIC

The Atherosclerosis Risk in Communities (ARIC) study is a population-based prospective cohort study of atherosclerosis and its sequelae. ARIC included 15,792 individuals, predominantly European American and African American, aged 45-64 years at baseline (1987-89), chosen by probability sampling from four US communities (northwest suburbs of Minneapolis, Minnesota; Washington County, Maryland; Forsyth County,

North Carolina; and Jackson, Mississippi). Cohort members completed three additional triennial follow-up examinations, a fifth exam in 2011-2013, a sixth exam in 2016-2017, a seventh exam in 2018-2019, an eighth exam in 2020, a ninth exam in 2021-2022, a tenth exam in 2023, and an eleventh exam is ongoing. The ARIC study has been described in detail previously (PMC8667593).

### CARDIA

The Coronary Artery Risk Development in Young Adults (CARDIA) Study is a study examining the development and determinants of clinical and subclinical cardiovascular disease and their risk factors. It began in 1985-6 with a group of 5115 black and white men and women aged 18-30 years. The participants were selected so that there would be approximately the same number of people in subgroups of race, gender, education (high school or less and more than high school) and age (18-24 and 25-30) in each of 4 centers: Birmingham, AL; Chicago, IL; Minneapolis, MN; and Oakland, CA. These same participants were asked to participate in follow-up examinations during 1987-1988 (Year 2), 1990-1991 (Year 5), 1992-1993 (Year 7), 1995-1996 (Year 10), 2000-2001 (Year 15), 2005-2006 (Year 20), 2010-2011 (Year 25), and 2015-2016 (Year 30). A majority of the group has been examined at each of the follow-up examinations (91%, 86%,81%, 79%, 74%, 72%, 72%, and 71%, respectively). While the specific aims of each examination have varied, data have been collected on a variety of factors believed to be related to heart disease. These include conditions with clear links to heart disease such as blood pressure, cholesterol and other lipids, and glucose. Data have also been collected on physical measurements such as weight and body composition as well as lifestyle factors such as dietary and exercise patterns, substance use (tobacco and alcohol), behavioral and psychological variables, medical and family history, and other chemistries (e.g., insulin). In addition, subclinical atherosclerosis has been measured via echocardiography during Years 5, 10, 25, and 30, a chest CT scan during Years 15, 20, and 25, an abdominal CT scan during Year 25, and carotid ultrasound during Year 20. A brain MRI was performed on a subset of participants at Years 25 and 30. The CARDIA cohort, born between 1955 and 1968, has been influenced substantially by the obesity epidemic at ages younger than participants in other established NHLBI cohorts. Further investigation of the mechanisms linking obesity to derangements in cardiovascular structure and function and the etiology of clinical events promises to generate important new knowledge to inform health promotion and disease prevention efforts.

### CHS

The Cardiovascular Health Study (CHS) is a population-based cohort study initiated by the National Heart, Lung and Blood Institute (NHLBI) in 1987 to determine the risk factors for development and progression of cardiovascular disease (CVD) in older adults, with an emphasis on subclinical measures. The study recruited 5,888 adults aged 65 or older at entry in four U.S. communities and conducted extensive annual clinical exams between 1989-1999 along with semi-annual phone calls, events adjudication, and subsequent data analyses and publications. Additional data are collected by studies ancillary to CHS. In June 1990, four Field Centers (Sacramento, CA; Hagerstown, MD; Winston-Salem, NC; Pittsburgh, PA) completed the recruitment of 5201 participants. Between November 1992 and June 1993, an additional 687 African Americans were recruited using similar methods. Blood samples were drawn from all participants at their baseline examination and during follow-up clinic visits and DNA was subsequently extracted from available samples. CHS analyses were limited to participants with available DNA who consented to genetic studies. The baseline examinations consisted of a home interview and a clinic examination that assessed not only traditional risk factors but also measures of subclinical disease, including carotid ultrasound, echocardiography, electrocardiography, and pulmonary function. Between enrollment and 1998-99, participants were seen in the clinic annually, and contacted by phone at 6-month intervals to collect information about hospitalizations and potential cardiovascular events. Major exam components were repeated during annual follow-up examinations through 1999. Cranial MRI scans, retinal photography, and tests of endothelial function were added as new components. Standard protocols for the identification and adjudication of events were implemented during follow-up. The adjudicated events are CHD, angina, heart failure (HF), stroke, transient ischemic attack (TIA), claudication and mortality. Adjudication of cause of death continues using a streamlined protocol; adjudication of other events ended in June 2015. Deep venous thrombosis and pulmonary embolism events from baseline through 2001 were adjudicated in an ancillary study: the Longitudinal Investigation of Thromboembolism Etiology (LITE). Since 1999, participants have been contacted every 6 months by phone, primarily to ascertain health status and for events follow-up. The study was initially approved by institutional review boards at the Field Centers (Wake Forest, University of California – Davis, Johns Hopkins University, University of Pittsburgh), the Core Laboratory (University of Vermont) and at the Coordinating Center (University of Washington). The University of Washington now handles CHS Data Repository approvals.

### FHS

The Framingham Heart Study (FHS) is a long-running, ongoing cardiovascular cohort study of residents of the city of Framingham, Massachusetts. FHS was established in 1948 to improve understanding of the common factors or characteristics that contribute to CVD. The first generation referred to as the Original cohort was recruited in 1948 (5209 mostly of European descent residents of the town of Framingham, MA, who have undergone biennial examinations since then),(11) the second generation referred to as the Offspring cohort was recruited in 1971 (5124 offspring of the original cohort and spouse of the Offspring who were examined every 4 to 8 years)(12) and the third generation referred to as Gen 3 (4095 children of the Offspring cohort enrolled in 2002 and examined periodically with a target interval of 4 years between examinations) was recruited between 2002 and 2005.(13) The first generation has been examined every two years. The second generation has been examined every 4-8 years. A small number of spouse individuals of the second generation were examined at the same time when the third generation had their first examination. To date, the survivors of all three generation cohorts are under active surveillance for cardiovascular events, stroke, and dementia.

### GeneSTAR

The Genetic Study of Atherosclerosis Risk (GeneSTAR) is an ongoing European- and African American-family-based study to investigate mechanisms of stroke and coronary heart disease. GeneSTAR was conducted initially in healthy adult siblings of probands with documented early onset coronary disease under 60 years of age from 1983-2007 in one of ten Baltimore, Maryland hospitals. Apparently healthy siblings of the probands and offspring of the siblings and probands were screened for traditional coronary disease and stroke risk factors from 2003-2006, during which time GeneSTAR enrolled 2,194 participants from 568 families who had no coronary disease, vascular thrombotic events, peripheral vascular disease, serious gastrointestinal disorders, autoimmune diseases, bleeding disorders, or hemorrhagic events. A subset of this study population participated in an MRI study between 2009 and 2013.

#### GENOA

The Genetic Epidemiology Network of Arteriopathy (GENOA) study, a part of the Family Blood Pressure Program (FBPP Investigators,2002), consists of hypertensive sibships that were recruited for linkage and association studies to identify genes that influence blood pressure and its target organ damage (Daniels, 2004). In the initial phase of the GENOA study (Phase I: 1996-2001), all members of sibships containing ≥ 2 individuals with essential hypertension clinically diagnosed before age 60 were invited to participate, including both hypertensive and normotensive siblings. In the second phase of the GENOA study (Phase II: 2000-2004), 1,239 non-Hispanic white and 1,482 African American participants were successfully re-recruited to measure potential target organ damage due to hypertension.

#### MESA

The Multi-Ethnic Study of Atherosclerosis (MESA) is a study of the characteristics of subclinical cardiovascular disease (disease detected non-invasively before it has produced clinical signs and symptoms) and the risk factors that predict progression to clinically overt cardiovascular disease or progression of the subclinical disease. MESA researchers study a diverse, population-based sample of 6,814 men and women aged 45-84 and free of clinically recognized cardiovascular disease. Thirty-eight percent of the recruited participants are White, 28 percent Black, 22 percent Hispanic, and 12 percent Asian, predominantly of Chinese descent. Participants were recruited from six field centers across the United States: Baltimore, Maryland; Chicago, Illinois; Forsyth County, North Carolina; Los Angeles County, California; New York, New York; and St. Paul, Minnesota. Participants are being followed for identification and characterization of cardiovascular disease events, including acute myocardial infarction and other forms of coronary heart disease (CHD), stroke, and heart failure; for cardiovascular disease interventions; and for mortality. In addition to the six Field Centers, MESA involves a Coordinating Center, a Central Laboratory, and Central Reading Centers for Computed Tomography (CT), Magnetic Resonance Imaging (MRI), Ultrasound, and Electrocardiography (ECG). The first examination took place over two years, from July 2000 - July 2002. It was followed by six examination periods that were 17-20 months in length. Participants have been contacted every 9 to 12 months throughout the study to assess clinical morbidity and mortality.

#### SAFHS

The San Antonio Family Heart Study (SAFHS) began in 1991 and included 1,431 individuals in 42 extended families at baseline. Probands were 40- to 60-year-old low-income Mexican Americans selected at random without regard to presence or absence of disease, almost exclusively from Mexican American census tracts in San Antonio, Texas. As part of ongoing studies, new family members were recruited to the original families, expanding the cohort to almost 3,099 individuals primarily from 73 families.

### Acknowledgements

#### TOPMed sequencing

Whole genome sequencing (WGS) for the Trans-Omics in Precision Medicine (TOPMed) program was supported by the National Heart, Lung and Blood Institute (NHLBI). Centralized read mapping and genotype calling, along with variant quality metrics and filtering were provided by the TOPMed Informatics Research Center (3R01HL-117626-02S1). Phenotype harmonization, data management, sample-identity QC, and general study coordination, were provided by the TOPMed Data Coordinating Center (3R01HL-120393-02S1). We gratefully acknowledge the studies and participants who provided biological samples and data for TOPMed. The Genome Sequencing Program (GSP) was funded by the National Human Genome Research Institute (NHGRI), the National Heart, Lung, and Blood Institute (NHLBI), and the National Eye Institute (NEI). The GSP Coordinating Center (U24 HG008956) contributed to cross program scientific initiatives and provides logistical and general study coordination. The Centers for Common Disease Genomics (CCDG) program was supported by NHGRI and NHLBI, and whole genome sequencing was performed at the Baylor College of Medicine Human Genome Sequencing Center (UM1 HG08898).

WGS for “NHLBI TOPMed: Atherosclerosis Risk in Communities (ARIC)” (phs001211.v4.p3) was performed at the Baylor College of Medicine Human Genome Sequencing Center (HHSN268201500015C and 3U54HG003273-12S2) and the Broad Institute for MIT and Harvard (3R01HL092577-06S1). WGS for “NHLBI TOPMed: Coronary Artery Risk Development in Young Adults (CARDIA)” (phs001612.v2.p2) was performed at Baylor College of Medicine Human Genome Sequencing Center (HHSN268201600033I). WGS for “NHLBI TOPMed: Cardiovascular Health Study (CHS)” (phs001368.v4.p2) was performed at Baylor College of Medicine Human Genome Sequencing Center (HHSN268201600033I). WGS for “NHLBI TOPMed: Whole Genome Sequencing and Related Phenotypes in the Framingham Heart Study (FHS)” (phs000974.v5.p4) was performed at the Broad Institute of MIT and Harvard (3R01HL092577-06S1, 3U54HG003067-12S2). WGS for “NHLBI TOPMed: Genetic Study of Atherosclerosis Risk (GeneSTAR)” (phs001218.v3.p1) was performed at Illumina (R01HL112064), Psomagen (formerly Macrogen, 3R01HL112064-04S1), and the Broad Institute of MIT and Harvard (HHSN268201500014C). WGS for “NHLBI TOPMed: Genetic Epidemiology Network of Arteriopathy (GENOA)” (phs001345.v3.p1) was performed at Northwest Genomics Center (3R01HL055673-18S1). WGS for “NHLBI TOPMed: Multi-Ethnic Study of Atherosclerosis (MESA)” (phs001416.v3.p1) was performed at the Broad Institute of MIT and Harvard (3U54HG003067-13S1). WGS for “NHLBI TOPMed: San Antonio Family Heart Study (SAFHS)” (phs001215.v4,p2) was performed at Illumina (3R01HL113323-03S1, R01HL113323).

### Study-specific data and Study ethic statements

#### NHLBI TOPMed: Atherosclerosis Risk in Communities Study VTE cohort (ARIC)

The Atherosclerosis Risk in Communities study has been funded in whole or in part with Federal funds from the National Heart, Lung, and Blood Institute, National Institutes of Health, Department of Health and Human Services, under Contract nos. (75N92022D00001, 75N92022D00002, 75N92022D00003, 75N92022D00004, 75N92022D00005). The authors thank the staff and participants of the ARIC study for their important contributions. All ARIC participants provided written informed consent, and the study design and methods were approved by the Institutional Review Boards at each of the collaborating medical centers: University of Mississippi Medical Center (Jackson Field Center); Wake Forest University Health Sciences (Forsyth County Field Center); University of Minnesota (Minnesota Field Center); and Johns Hopkins University School of Public Health (Washington County Field Center).

#### NHLBI TOPMed: Coronary Artery Risk Development in Young Adults (CARDIA)

The Coronary Artery Risk Development in Young Adults Study (CARDIA) is conducted and supported by the National Heart, Lung, and Blood Institute (NHLBI) in collaboration with the University of Alabama at

Birmingham (75N92023D00002 & 75N92023D00005), Northwestern University (75N92023D00004), University of Minnesota (75N92023D00006), and Kaiser Foundation Research Institute (75N92023D00003). CARDIA was also partially supported by the Intramural Research Program of the National Institute on Aging (NIA) and an intra-agency agreement between NIA and NHLBI (AG0005). All CARDIA participants provided written informed consent, and the study was approved by the Institutional Review Boards of the participating institutions: Kaiser Permanente Division of Research (Oakland Field Center), Northwestern University (Chicago Field Center), University of Minnesota (Minneapolis Field Center), University of Alabama at Birmingham (Birmingham Field center and Coordinating Center), the University of Texas Health Science Center at Houston (DNA laboratory).

#### NHLBI TOPMed: Cardiovascular Health Study (CHS)

Cardiovascular Health Study: This research was supported by contracts HHSN268201200036C, HHSN268200800007C, HHSN268201800001C, N01HC55222, N01HC85079, N01HC85080, N01HC85081, N01HC85082, N01HC85083, N01HC85086, N01HC15103, 75N92021D00006 and grants R01HL105756, U01HL080295, U01HL130114, and R01HL172803 from the National Heart, Lung, and Blood Institute (NHLBI), with additional contribution from the National Institute of Neurological Disorders and Stroke (NINDS). Additional support was provided by R01AG023629, R01AG15928, R01AG20098 from the National Institute on Aging (NIA). A full list of principal CHS investigators and institutions can be found at CHS-NHLBI.org. The content is solely the responsibility of the authors and does not necessarily represent the official views of the National Institutes of Health. All CHS participants provided informed consent, and the study was approved by the Institutional Review Board of University Washington.

#### NHLBI TOPMed: Framingham Heart Study (FHS)

The Framingham Heart Study is conducted and supported by the National Heart, Lung, and Blood Institute (NHLBI) in collaboration with Boston University (Contract No. N01-HC-25195, HHSN268201500001I and 75N92019D00031). This work was also supported by grant R01AG016495, R01AG063507, R01AG054076, R01AG049607, R01AG059421, R01AG033040, R01AG066524, P30AG066546, U01 AG052409, U01 AG058589 from the National Institute on Aging and R01 AG017950, UH2/3 NS100605, UF1 NS125513 from National Institute of Neurological Disorders and Stroke and R01HL132320. We also acknowledge the dedication of the FHS study participants without whom this research would not be possible. The Framingham Heart Study was approved by the Institutional Review Board of the Boston University Medical Center. All study participants provided written informed consent.

#### NHLBI TOPMed: Genetic Studies of Atherosclerosis Risk (GeneSTAR)

GeneSTAR was supported by the National Institutes of Health/National Heart, Lung, and Blood Institute (U01 HL72518, HL087698, HL112064), the National Institutes of Health/National Institute of Neurological Disorders and Stroke (NS062059), and by a grant from the National Institutes of Health/National Center for Research Resources (M01-RR000052) to the Johns Hopkins General Clinical Research Center, and the National Center for Research Resources and the National Center for Advancing Translational Sciences, National Institutes of Health (UL1 RR 025005) to the Johns Hopkins Institute for Clinical & Translational Research. We would like to thank the participants and families of GeneSTAR and our dedicated staff for all their sacrifices. All participants provided written informed consent and the study was approved by the Johns Hopkins Medicine Institutional Review Board.

#### NHLBI TOPMed: Genetic Epidemiology Network of Arteriopathy (GENOA)

Support for GENOA was provided by the National Heart, Lung and Blood Institute (U01HL054457, U01HL054464, U01HL054481, R01HL119443, and R01HL087660) and the National Institute of Neurological Disorders and Stroke (R01NS041558) of the National Institutes of Health. Whole-genome sequencing of GENOA was supported by R01 HL055673-18S1. Written informed consent was obtained from all subjects and approval was granted by participating institutional review boards (University of Michigan, University of Mississippi Medical Center, and Mayo Clinic).

#### NHLBI TOPMed: Multi-Ethnic Study of Atherosclerosis (MESA)

Whole genome sequencing (WGS) for the Trans-Omics in Precision Medicine (TOPMed) program was supported by the National Heart, Lung and Blood Institute (NHLBI). WGS for “NHLBI TOPMed: Multi-Ethnic Study of Atherosclerosis (MESA)” (phs001416.v3.p1) was performed at the Broad Institute of MIT and Harvard (3U54HG003067-13S1). Centralized read mapping and genotype calling, along with variant quality metrics and filtering were provided by the TOPMed Informatics Research Center (3R01HL-117626-02S1). Phenotype harmonization, data management, sample-identity QC, and general study coordination, were provided by the TOPMed Data Coordinating Center (3R01HL-120393-02S1), and TOPMed MESA Multi-Omics (HHSN2682015000031/HSN26800004). The MESA projects are conducted and supported by the National Heart, Lung, and Blood Institute (NHLBI) in collaboration with MESA investigators. Support for the Multi-Ethnic Study of Atherosclerosis (MESA) projects are conducted and supported by the National Heart, Lung, and Blood Institute (NHLBI) in collaboration with MESA investigators. Support for MESA is provided by contracts 75N92020D00001, HHSN268201500003I, N01-HC-95159, 75N92020D00005, N01-HC-95160, 75N92020D00002, N01-HC-95161, 75N92020D00003, N01-HC-95162, 75N92020D00006, N01-HC-95163, 75N92020D00004, N01-HC-95164, 75N92020D00007, N01-HC-95165, N01-HC-95166, N01-HC-95167, N01-HC-95168, N01-HC-95169, UL1-TR-000040, UL1-TR-001079, UL1-TR-001420, UL1TR001881, DK063491, R01HL105756, and R01HL127659. The authors thank the other investigators, the staff, and the participants of the MESA study for their valuable contributions. A full list of participating MESA investigators and institutes can be found at http://www.mesa-nhlbi.org.

#### NHLBI TOPMed: Whole Genome Sequencing to Identify Causal Genetic Variants Influencing CVD Risk - San Antonio Family Studies (SAFHS)

Collection of the San Antonio Family Study data was supported in part by National Institutes of Health (NIH) grants R01 HL045522, MH078143, MH078111 and MH083824; and whole genome sequencing of SAFHS subjects was supported by U01 DK085524 and R01 HL113323. We are very grateful to the participants of the San Antonio Family Study for their continued involvement in our research programs. All SAFHS participants provided informed consent, and the study was approved by the Institutional Review Board at the University of Texas Rio Grande Valley.

### Supplementary Tables

#### **Supplementary Table 1:** MRI scanner, protocol, and volumetric quantification approach by study

| **Study** | MRI-scanner (Tesla) | MRI protocol used | Method for quantifying brain volumes |
|---|---|---|---|
| **ARIC** | Structural brain images were obtained using 3-T MRI scanners (Siemens Verio [Maryland study center], Siemens Skyra [North Carolina and Mississippi study centers], or Siemens Trio [Minnesota study center] following identical protocols (PMID: 28916531). | The scans included sagittal T1-weighted magnetization–prepared rapid gradient-echo (MPRAGE) images (1.2-mm slices), axial gradient recalled echo T2-weighted imaging (T2*GRE) (4-mm slices), axial T2 fluid-attenuated inversion recovery (FLAIR) (5-mm slices), field mapping (3-mm slices), and axial diffusion tensor images (2.7-mm slices for Skyra and Verio scanners and 3-mm slices for Trio scanner). Brain volumes were estimated based on T1-weighted scans using image analysis software (FreeSurfer; http://surfer.nmr.mgh.harvard.edu) (PMID: 11832223). Image processing was done at the ARIC MRI Reading Center at the Mayo Clinic (Rochester, MN) according to methods developed for ADNI (PMID: 20451869). | Freesurfer v5.3 |
| **CARDIA** | 3T Siemens TIM Trio, 3T Philips Achieva | We used the following sequence for morphological analysis: Sagital 3D T1 MPRAGE (Tr 1900 Te 2.89 Fov 250mm, thickness 1mm slices 176 slices, Base Res 255, Phase res 100%, Matrix 256X256 NSA 1 TI 900 ms Pixel BW 170hz. ETL = 1 Flip = 9). We estimated WMH from sagittal 3D FLAIR (Tr 6000 Te 160 Fov 250mm (fov phase = 85%), thickness 1 mm slices 160 slices, Base Res 202, Phase res 91%, Matrix 258 X 221 NSA 1 TI = 2200 ms, Pixel BW 930, ETL 203)), T1 and T2 (Plane Sagittal Coil 12 channel Psd File name 3D T2: Tr 3200 Te 409 Fov 250 (fov phase = 80%), thickness 1mm slices 176, Base Res = 246, Phase res = 80%, Matrix 258x256 NSA 1 Center Freq. water ETL 141 Flip 120 Pixel BW 750)] sequences. Total intracranial volume was measured as the sum of gray matter, white matter, and cerebral spinal fluid (CSF) volumes. | Automated adaptive Bayesian segmentation and HAMMER or RAVENS |
| **CHS** | 1.5T GE or Picker | The scans included sagittal T1-weighted magnetization–prepared rapid gradient-echo (MPRAGE) imaging, axial T2 fluid attenuation inversion recovery (FLAIR), and axial DTI pulse sequences. Brain volumes were estimated based on T1-weighted scans. | FreeSurfer v 5.1 (prospective) |
| **FHS** | The MRI protocol in FHS has been described previously [PMID: 14716734]. Briefly, participants were imaged by a variety of MRI machines varying in field strength from 1 to 3 Tesla. | Two sequences were used: a 3-dimensional T1-weighted and 2 or 3-dimensional FLAIR imaging. All images were transferred to and processed by the University of California Davis Medical Center without knowledge of clinical information. Segmentation and quantification of brain volume measures were performed by automated procedures with quality control. Total cerebral cranial volume (TCV) was determined using a convolutional neural network method [PMID: 34234642]. Images were further segmented into 4 tissue types (gray, white, CSF, and WMH volumes) using previously published methods[PMID: 23367130, PMID: 35382232, PMID: 23365843]. Hippocampal analyses were performed using atlas based [PMID: 25223727] diffeomorphic approach [PMID: 17354834] with the minor modification of label refinement. Non-linear co-registration of images to the DKT atlas [PMID: 16530430] enabled calculation of regional gray matter volumes [PMID: 17354834, PMID: 16530430, PMID: 19245840]. | In-house protocol and Atlas-based approach |

| | | | |
|---|---|---|---|
| **GeneSTAR** | 3T Philips Achieva | MPRAGE images were skull- stripped and co- registered to FLAIR images. Spatial normalization of the co-registered MPRAGE and FLAIR images into MNI space was performed via affine transformation. Following sequences were used for WMH quantification: 1) Axial T1-weighted MPRAGE (magnetization prepared rapid gradient echo): TR (repetition time) 10 ms, TE (time to echo) 6 ms, TI (inversion time) 983 ms, voxel size 0.75 × 0.75 × 1.0 mm3, contiguous slices, with field of view imaging (FOV) 240 mm, matrix 256×256×160 mm. 2) Axial turbo spin echo FLAIR (fluid attenuated inversion recovery): TR 11000 ms, TI 2800 ms, TE 68 ms, voxel size 0.47 × 0.47 × 3.0 mm3, contiguous slices, FOV 240 mm, matrix 256 X 256 mm. | MIPAV (NIH, U-Penn) |
| **GENOA** | 1.5T GE Signa | Total intracranial volume (head size) was measured from T1-weighted spin echo sagittal images, each set consisting of 32 contiguous 5 mm thick slices with no interslice gap, field of view = 24 cm, matrix = 256 x 192, obtained with the following sequence: scan time = 2.5 min, echo time = 14 ms, repetitions = 2, replication time = 500 ms5. Total brain and leukoaraiosis volumes were determined from axial fluid-attenuated inversion recovery (FLAIR) images, each set consisting of 48 contiguous 3-mm interleaved slices with no interslice gap, field of view = 22 cm, matrix = 256 x 160, obtained with the following sequence: scan time = 9 min, echo time = 144.8 ms, inversion time = 2,600 ms, repetition time = 26,002 ms, bandwidth = +/- 15.6 kHz, one signal average. A fully automated algorithm was used to segment each slice of the edited multi-slice FLAIR sequence into voxels assigned to one of three categories: brain, cerebrospinal fluid, or leukoaraiosis | In-house protocol |
| **MESA** | 3T Siemens Prisma or Skyra | Imaging parameters were as follows: T1 (TR=1900ms, TE=2.93ms, flip angle=9, FOV=250mm, slice thickness=1mm, slices=176, averages=1); T2 (TR=3200ms, TE=408ms, FOV=250mm, slice thickness=1mm, slices=176, averages=1); FLAIR (TR=6000ms, TE=289ms, TI=2200 ms, FOV= 258mm, slice thickness=1mm, slices=160) | Multi-atlas based segmentation |
| **SAFHS** | 3T Siemens TIM Trio | The protocol included seven high-resolution T1- weighted 3D turbo-flash sequences with an adiabatic inversion contrast pulse and the following parameters: TE/TR/TI = 3.04/2100/785 ms, flip angle = 13°, 0.8 mm isotropic resolution, 200 mm FOV, 5-min duration (35-min total). A single image was obtained by linearly co- registering these images and computing the average, allowing improvement over the signal-to-noise ratio and reducing motion artifacts | FreeSurfer v 5.1.0 |

**Supplementary Table 2**: Differences in sub-volumes used in phenotype construction among TOPMed studies

| Study | ICV includes infratentorial volume | TBV includes infratentorial volume | TBV includes ventricular volumes |
|---|---|---|---|
| FHS | Yes | Yes | Yes |
| MESA | Yes | Yes | No |
| GeneSTAR | Yes | No | Yes |
| SAFHS | Yes | Yes | Yes |
| ARIC | Yes | Yes | No |

| | | | |
|---|---|---|---|
| CARDIA | Yes | Yes | No |
| CHS | Yes | Yes | No |
| GENOA | Yes | Yes | Yes |

ICV = intracranial volume; TBV = total brain volume.

**Supplementary Table 3**: Lookup of known brain volume loci in TOPMed brain MRI WGS association analysis

| Chromosome | Position (hg38) | Freq | MAC | Effect/ Non-effect alleles | Effect | P-value |
|---|---|---|---|---|---|---|
| **Hippocampal Volume (N=7,040)** | | | | | | |
| 1 | 48,043,347 | 0.592 | 5,750 | G/T | 0.0165 | 0.17 |
| 1 | 48,048,827 | 0.592 | 5,740 | T/C | 0.0134 | 0.27 |
| 1 | 54,389,311 | 0.568 | 6,078 | A/G | -0.0361 | 0.003 |
| 1 | 153,884,022 | 0.543 | 6,428 | T/C | 0.0204 | 0.086 |
| 2 | 161,057,657 | 0.251 | 3,537 | A/G | -0.0271 | 0.048 |
| 2 | 161,989,055 | 0.630 | 5,214 | C/T | -0.0121 | 0.33 |
| 2 | 161,999,638 | 0.545 | 6,403 | T/C | -0.0038 | 0.75 |
| 3 | 142,040,538 | 0.748 | 3,547 | T/C | -0.034 | 0.012 |
| 3 | 171,318,238 | 0.115 | 1,619 | G/C | 0.0355 | 0.052 |
| 3 | 171,375,102 | 0.302 | 4,248 | T/A | 0.0231 | 0.074 |
| 3 | 190,925,050 | 0.360 | 5,069 | A/G | 0.0285 | 0.02 |
| 4 | 184,034,308 | 0.387 | 5,453 | A/T | -0.0299 | 0.013 |
| 5 | 66,729,141 | 0.744 | 3,606 | A/G | -0.0366 | 0.0075 |
| 5 | 66,785,101 | 0.400 | 5,631 | T/C | -0.0385 | 0.0014 |
| 5 | 66,788,432 | 0.345 | 4,862 | T/G | -0.0334 | 0.0068 |
| 5 | 66,816,887 | 0.265 | 3,727 | A/G | 0.0476 | 0.0004 |
| 5 | 83,562,195 | 0.151 | 2,133 | G/C | 0.0368 | 0.022 |
| 6 | 108,676,925 | 0.123 | 1,734 | G/T | -0.0134 | 0.44 |
| 6 | 149,613,041 | 0.437 | 6,148 | A/G | 0.0482 | 9.70E-05 |
| 7 | 31,390,884 | 0.145 | 2,038 | G/C | -0.03 | 0.069 |
| 7 | 31,406,487 | 0.160 | 2,251 | G/A | -0.025 | 0.12 |
| 7 | 149,155,342 | 0.575 | 5,987 | A/C | -0.0241 | 0.058 |
| 7 | 156,013,260 | 0.247 | 3,472 | T/A | 0.0276 | 0.041 |
| 7 | 156,014,259 | 0.247 | 3,476 | T/A | 0.0275 | 0.042 |
| 9 | 93,323,088 | 0.466 | 6,568 | T/C | 0.0057 | 0.65 |
| 9 | 116,482,848 | 0.211 | 2,969 | C/T | -0.0312 | 0.029 |
| 9 | 116,482,904 | 0.343 | 4,834 | A/G | 0.0368 | 0.0038 |
| 9 | 116,485,695 | 0.370 | 5,207 | C/G | 0.0377 | 0.0024 |
| 9 | 116,498,079 | 0.577 | 5,961 | G/C | 0.0135 | 0.26 |
| 10 | 124,735,568 | 0.522 | 6,729 | T/C | -0.0341 | 0.0055 |
| 10 | 124,742,712 | 0.536 | 6,536 | C/G | -0.0325 | 0.0074 |
| 10 | 124,785,631 | 0.557 | 6,239 | T/C | -0.0262 | 0.029 |
| 12 | 3,898,158 | 0.418 | 5,879 | G/T | -0.0147 | 0.23 |

| Chromosome | Position (hg38) | Freq | MAC | Effect/ Non-effect alleles | Effect | P-value |
|---|---|---|---|---|---|---|
| 12 | 3,898,732 | 0.488 | 6,876 | C/T | 0.0136 | 0.27 |
| 12 | 65,360,462 | 0.240 | 3,383 | A/C | -0.0326 | 0.02 |
| 12 | 65,372,164 | 0.208 | 2,931 | A/G | -0.0371 | 0.012 |
| 12 | 65,402,530 | 0.188 | 2,653 | A/T | 0.0126 | 0.4 |
| 12 | 65,438,688 | 0.119 | 1,669 | G/T | -0.0507 | 0.0048 |
| 12 | 65,550,385 | 0.241 | 3,395 | G/T | -0.0244 | 0.1 |
| 12 | 116,825,755 | 0.620 | 5,352 | A/G | 0.0501 | 0.00014 |
| 12 | 116,849,176 | 0.643 | 5,027 | T/C | 0.0481 | 0.00037 |
| 12 | 116,883,857 | 0.118 | 1,666 | C/T | 0.0798 | 1.00E-05 |
| 12 | 116,885,562 | 0.077 | 1,088 | C/T | 0.106 | 8.30E-07 |
| 12 | 116,975,545 | 0.751 | 3,512 | G/A | -0.0136 | 0.38 |
| 15 | 97,899,241 | 0.705 | 4,158 | G/C | -0.0259 | 0.047 |
| 16 | 30,091,839 | 0.435 | 6,130 | A/C | 0.0129 | 0.28 |
| 16 | 30,114,519 | 0.541 | 6,463 | C/G | 0.0035 | 0.77 |
| 19 | 44,908,684 | 0.141 | 1,988 | C/T | -0.021 | 0.22 |
| **Intracranial Volume (N=7,673)** | | | | | | |
| 3 | 190,953,113 | 0.049 | 757 | C/G | 11.4 | 0.006 |
| 6 | 108,624,167 | 0.587 | 6,334 | C/A | 11.2 | 6.70E-09 |
| 6 | 126,470,949 | 0.252 | 3,867 | G/A | 10.2 | 3.30E-06 |
| 10 | 103,410,892 | 0.275 | 4,224 | T/G | 4.2 | 0.047 |
| 12 | 65,980,467 | 0.571 | 6,579 | G/A | -5.5 | 0.0031 |
| 12 | 102,529,208 | 0.186 | 2,847 | C/G | -5.7 | 0.015 |
| 17 | 46,770,468 | 0.156 | 2,389 | G/T | -12.1 | 3.30E-06 |
| **Lateral Ventricular Volume (N=7,632)** | | | | | | |
| 3 | 190,902,357 | 0.347 | 5,294 | A/G | -0.032 | 5.80E-06 |
| 10 | 21,589,215 | 0.243 | 3,711 | A/T | -0.02 | 0.01 |
| 11 | 111,207,001 | 0.316 | 4,816 | G/A | -0.023 | 0.0013 |
| 12 | 106,083,027 | 0.067 | 1,025 | C/T | 0.072 | 4.00E-08 |
| 16 | 87,191,495 | 0.533 | 7,130 | A/G | 0.03 | 1.30E-05 |
| 22 | 37,714,443 | 0.553 | 6,816 | T/C | -0.019 | 0.0054 |
| **Total brain volume (N=7,604)** | | | | | | |
| 1 | 243,568,588 | 0.213 | 3,240 | C/T | 1.328 | 0.066 |
| 2 | 182,838,608 | 0.098 | 1,488 | A/G | -1.997 | 0.042 |
| 3 | 141,998,599 | 0.118 | 1,794 | G/A | -0.63 | 0.48 |
| 6 | 108,613,258 | 0.559 | 6,702 | T/C | 1.361 | 0.033 |
| 6 | 126,643,364 | 0.273 | 4,152 | G/A | -0.064 | 0.93 |
| 7 | 92,610,217 | 0.221 | 3,358 | G/A | -1.051 | 0.13 |
| 10 | 103,253,237 | 0.175 | 2,660 | C/T | 0.546 | 0.47 |
| 12 | 56,080,024 | 0.606 | 5,991 | C/T | -0.568 | 0.35 |
| 12 | 65,982,311 | 0.575 | 6,463 | C/T | 0.815 | 0.17 |
| 17 | 45,826,180 | 0.166 | 2,529 | G/A | -2.298 | 0.0036 |

MAC: minor allele count

**Supplementary Table 4**: Top 10 genes from gene-based rare variant association analyses of TBV in TOPMed

| gene | N variants | N | cMAC | min.pval | opt.rho | pval_SKATO |
|---|---|---|---|---|---|---|
| *PXK* | 53 | 233 | 252 | 7.9E-05 | 0.9 | 1.7E-04 |
| *CDKL3* | 51 | 337 | 347 | 9.7E-05 | 0.9 | 2.1E-04 |
| *SH2B2* | 78 | 338 | 355 | 1.4E-04 | 0.2 | 3.7E-04 |
| *GSX1* | 19 | 30 | 30 | 1.7E-04 | 0.9 | 4.2E-04 |
| *KAT5* | 21 | 179 | 179 | 1.8E-04 | 0 | 4.5E-04 |
| *KARS* | 60 | 225 | 229 | 2.5E-04 | 0.5 | 6.3E-04 |
| *IGKV2-28* | 18 | 26 | 29 | 6.1E-04 | 0.2 | 6.8E-04 |
| *CCDC9* | 87 | 345 | 357 | 3.4E-04 | 0 | 7.8E-04 |
| *SALL3* | 149 | 548 | 582 | 3.7E-04 | 0.9 | 8.6E-04 |
| *ARHGAP19-SLIT1* | 36 | 149 | 149 | 3.4E-04 | 0.1 | 9.0E-04 |

cMAC: cumulative minor allele count

A total of 19,534 genes were analyzed using SKAT-O for TBV.

min.pval: the minimum p-value among the p-values calculated for each choice of rho.

opt.rho: the optimal rho value; i.e. the rho value that gave the minimum p-value.

pval_SKATO: the SKAT-O p-value after adjustment for searching across multiple rho values.

**Supplementary Table 5**: Top 10 genes from gene-based rare variant association analyses of LVV in TOPMed

| gene | N variants | N | cMAC | min.pval | opt.rho | pval_SKATO |
|---|---|---|---|---|---|---|
| *RIPK1* | 45 | 106 | 106 | 5.0E-06 | 0.1 | 2.0E-05 |
| *CMTR1* | 48 | 107 | 109 | 1.8E-05 | 0 | 5.8E-05 |
| *IFITM2* | 28 | 99 | 101 | 2.9E-05 | 0.2 | 8.7E-05 |
| *XXYLT1* | 64 | 261 | 268 | 5.4E-05 | 0 | 1.5E-04 |
| *SLC10A2* | 51 | 282 | 284 | 8.5E-05 | 0 | 2.4E-04 |
| *PAK1* | 16 | 35 | 35 | 9.7E-05 | 0.9 | 2.6E-04 |
| *COL25A1* | 60 | 268 | 281 | 1.1E-04 | 0 | 2.8E-04 |
| *CHST7* | 18 | 112 | 139 | 1.2E-04 | 0 | 3.1E-04 |
| *H2AFB1* | 6 | 12 | 15 | 1.8E-04 | 1 | 3.8E-04 |
| *AP1AR* | 21 | 208 | 210 | 1.4E-04 | 0.3 | 3.8E-04 |

cMAC: cumulative minor allele count

A total of 19,523 genes were analyzed using SKAT-O for LVV.

min.pval: the minimum p-value among the p-values calculated for each choice of rho.

opt.rho: the optimal rho value; i.e. the rho value that gave the minimum p-value.

pval_SKATO: the SKAT-O p-value after adjustment for searching across multiple rho values.

**Supplementary Table 6**: Top 10 genes from gene-based rare variant association analyses of HV in TOPMed

| gene | N variants | N | cMAC | min.pval | opt.rho | pval_SKATO |
|---|---|---|---|---|---|---|
| *IFIT1* | 38 | 204 | 206 | 1.6E-05 | 0.1 | 5.4E-05 |
| *DEPTOR* | 36 | 214 | 216 | 1.5E-04 | 0 | 4.0E-04 |
| *INTS13* | 29 | 130 | 131 | 2.4E-04 | 0.9 | 6.2E-04 |
| *RASSF9* | 41 | 217 | 225 | 2.4E-04 | 0 | 6.2E-04 |
| *RFPL4A* | 42 | 204 | 213 | 2.5E-04 | 0.1 | 6.3E-04 |
| *DDX42* | 37 | 179 | 179 | 3.0E-04 | 0 | 7.8E-04 |
| *ANKRD34A* | 19 | 33 | 33 | 3.8E-04 | 0.9 | 9.1E-04 |
| *CELF4* | 36 | 173 | 176 | 3.5E-04 | 0.9 | 9.3E-04 |
| *PPIAL4C* | 24 | 269 | 285 | 3.8E-04 | 0.2 | 9.3E-04 |
| *TINF2* | 34 | 164 | 165 | 4.1E-04 | 0 | 1.0E-03 |

cMAC: cumulative minor allele count

A total of 19,344 genes were analyzed using SKAT-O for HV.

min.pval: the minimum p-value among the p-values calculated for each choice of rho.

opt.rho: the optimal rho value; i.e. the rho value that gave the minimum p-value.

pval_SKATO: the SKAT-O p-value after adjustment for searching across multiple rho values.

**Supplementary Table 7**: Top 10 genes from gene-based rare variant association analyses of ICV in TOPMed

| gene | N variants | N | cMAC | min.pval | opt.rho | pval_SKATO |
|---|---|---|---|---|---|---|
| *BIRC6* | 272 | 1140 | 1323 | 1.8E-05 | 0.9 | 4.6E-05 |
| *ZNF41* | 53 | 363 | 475 | 5.2E-05 | 0 | 1.4E-04 |
| *LAT2* | 23 | 68 | 68 | 5.3E-05 | 0 | 1.6E-04 |
| *NXT1* | 4 | 13 | 13 | 8.1E-05 | 1 | 1.8E-04 |
| *FAM163A* | 13 | 39 | 39 | 1.4E-04 | 0.2 | 3.6E-04 |
| *WDR48* | 18 | 81 | 81 | 1.4E-04 | 0 | 3.7E-04 |
| *OLIG1* | 22 | 128 | 130 | 1.5E-04 | 0 | 4.0E-04 |

| | | | | | | |
|---|---|---|---|---|---|---|
| *IGSF9B* | 114 | 497 | 513 | 2.0E-04 | 0 | 5.0E-04 |
| *COX6A1* | 16 | 25 | 25 | 2.7E-04 | 0 | 6.8E-04 |
| *STATH* | 11 | 40 | 40 | 3.1E-04 | 0 | 7.5E-04 |

cMAC: cumulative minor allele count

A total of 19,554 genes were analyzed using SKAT-O for ICV.

min.pval: the minimum p-value among the p-values calculated for each choice of rho.

opt.rho: the optimal rho value; i.e. the rho value that gave the minimum p-value.

pval_SKATO: the SKAT-O p-value after adjustment for searching across multiple rho values.

Supplementary Figures

**Supplementary Figure 1**: Principal components 1 and 2 for TOPMed participants included in the brain volume WGS association analysis by self-reported race or ethnicity

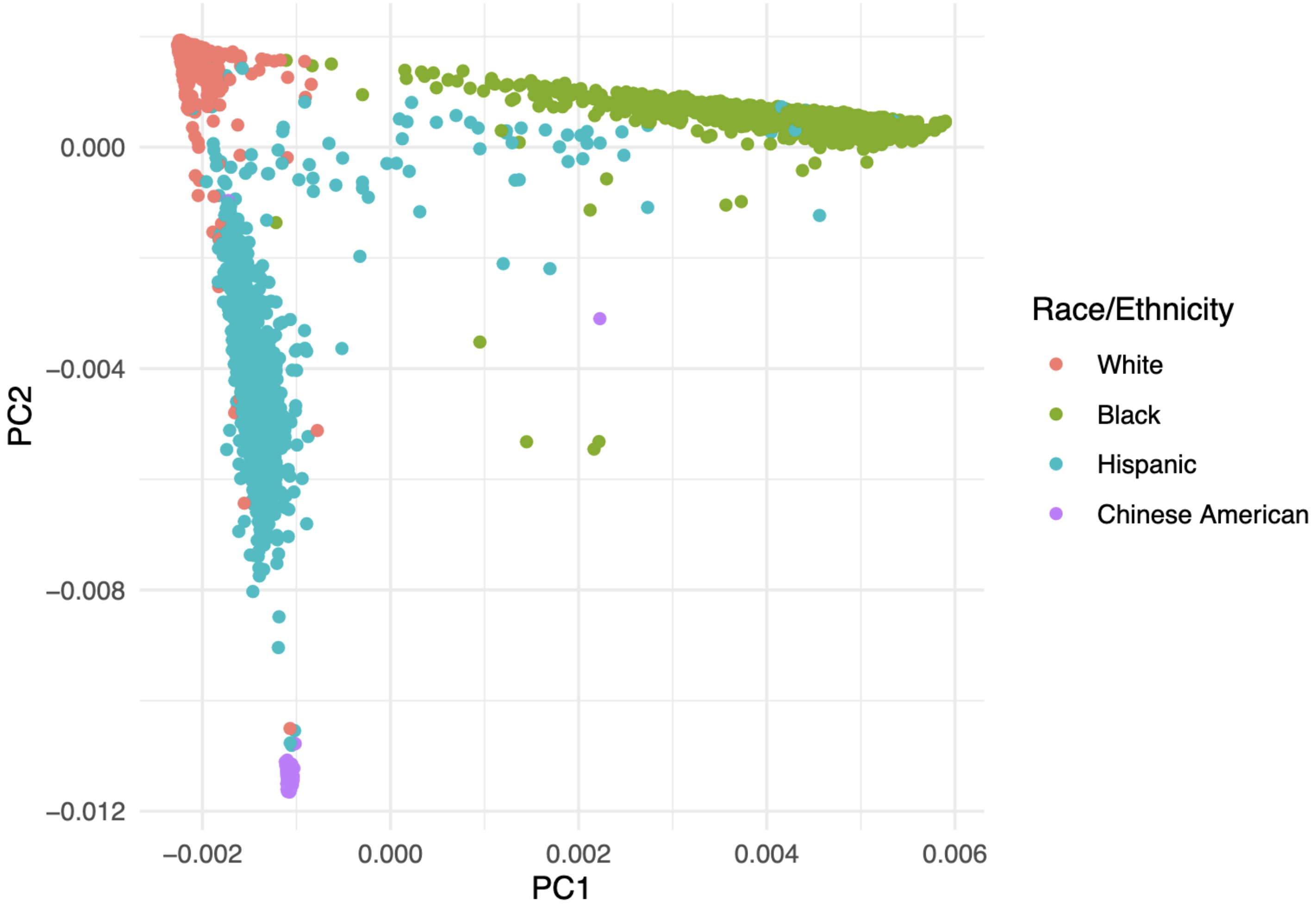

**Supplementary Figure 2**: QQ-plot for single variant association analysis of brain MRI volumes in TOPMed

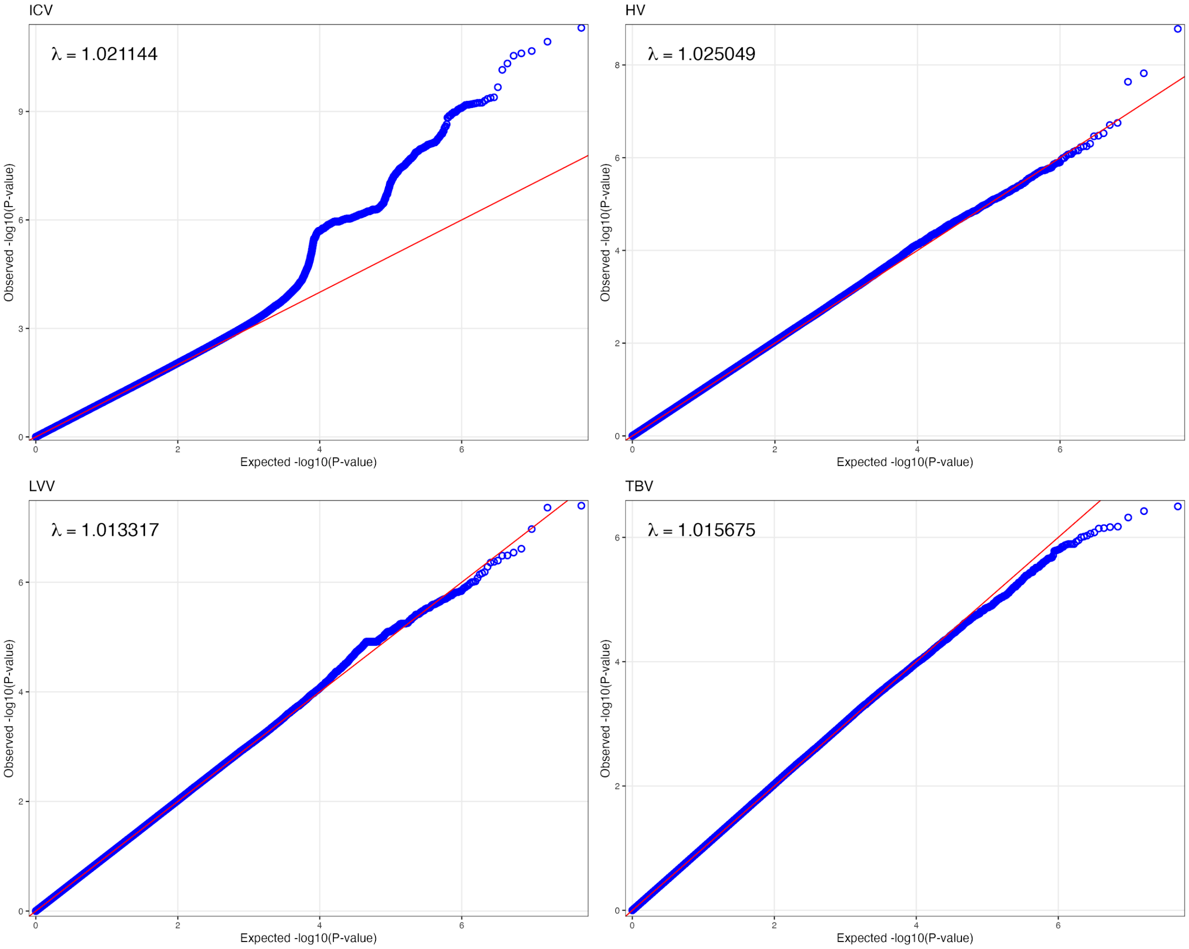


HV = hippocampal volume; ICV = intracranial volume; LVV = log lateral ventricular volume; TBV = total brain volume.

**Supplementary Figure 3**: Manhattan plots for association analysis of brain volumes in Black TOPMed participants

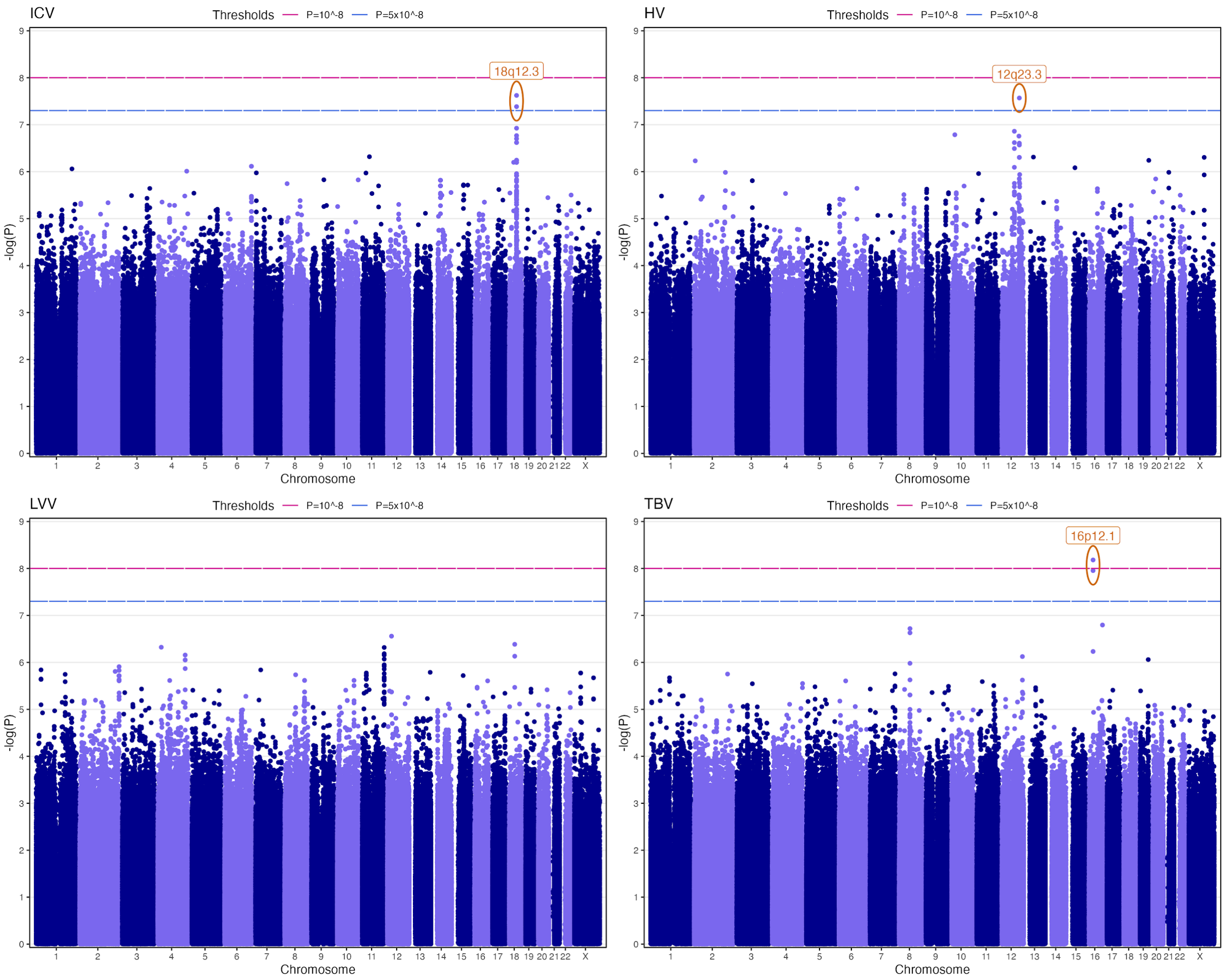


Blue dashed line demarcates sub-genome-wide significance ($P \leq 5x10^{-8}$). Pink dashed line demarcates genome-wide significance ($P \leq 10^{-8}$). HV = hippocampal volume; ICV = intracranial volume; LVV = log lateral ventricular volume; TBV = total brain volume. Only genetic variants associations passing minor allele count (MAC) ≥ 20 are represented.

**Supplementary Figure 4**: Manhattan plots for association analysis of brain volumes in Hispanic TOPMed participants

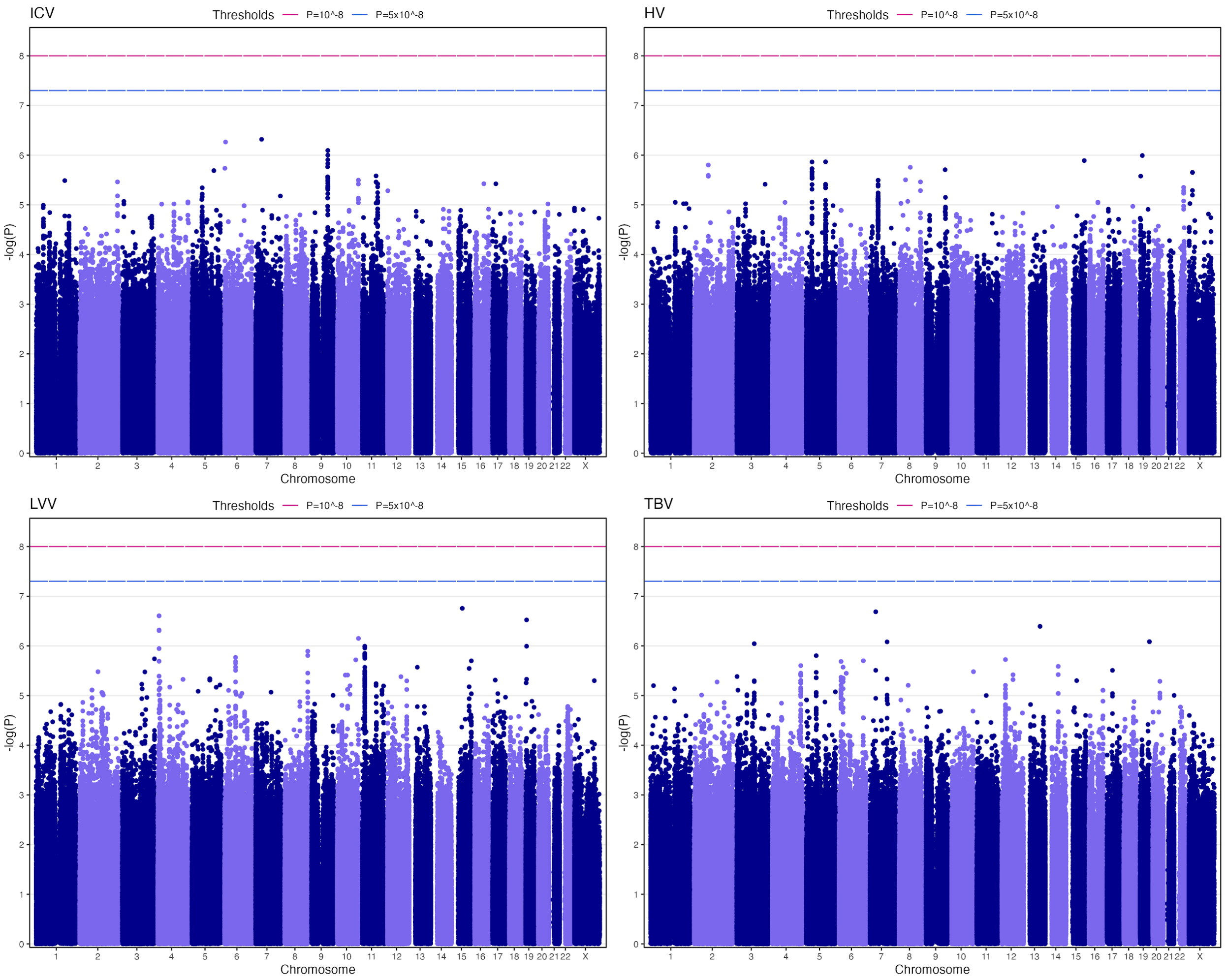


Blue dashed line demarcates sub-genome-wide significance ($P \leq 5x10^{-8}$). Pink dashed line demarcates genome-wide significance ($P \leq 10^{-8}$). HV = hippocampal volume; ICV = intracranial volume; LVV = log lateral ventricular volume; TBV = total brain volume. Only genetic variants associations passing minor allele count (MAC) ≥ 20 are represented.

**Supplementary Figure 5**: Manhattan plots for association analysis of brain volumes in White TOPMed participants

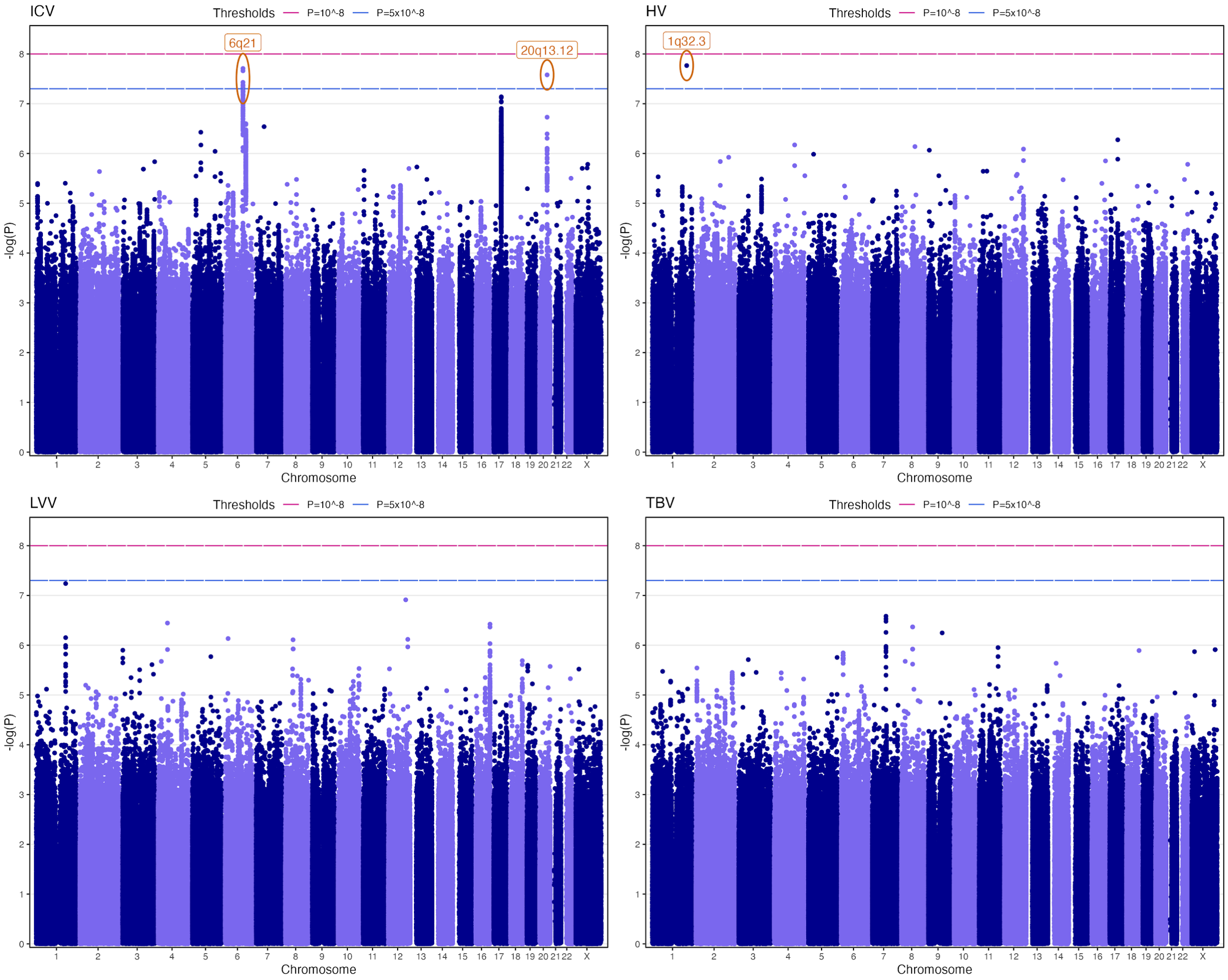


Blue dashed line demarcates sub-genome-wide significance ($P \leq 5x10^{-8}$). Pink dashed line demarcates genome-wide significance ($P \leq 10^{-8}$). HV = hippocampal volume; ICV = intracranial volume; LVV = log lateral ventricular volume; TBV = total brain volume. Only genetic variants associations passing minor allele count (MAC) ≥ 20 are represented.

**Supplementary Figure 6**: Regional association plots for ICV chr6 (pooled analysis)

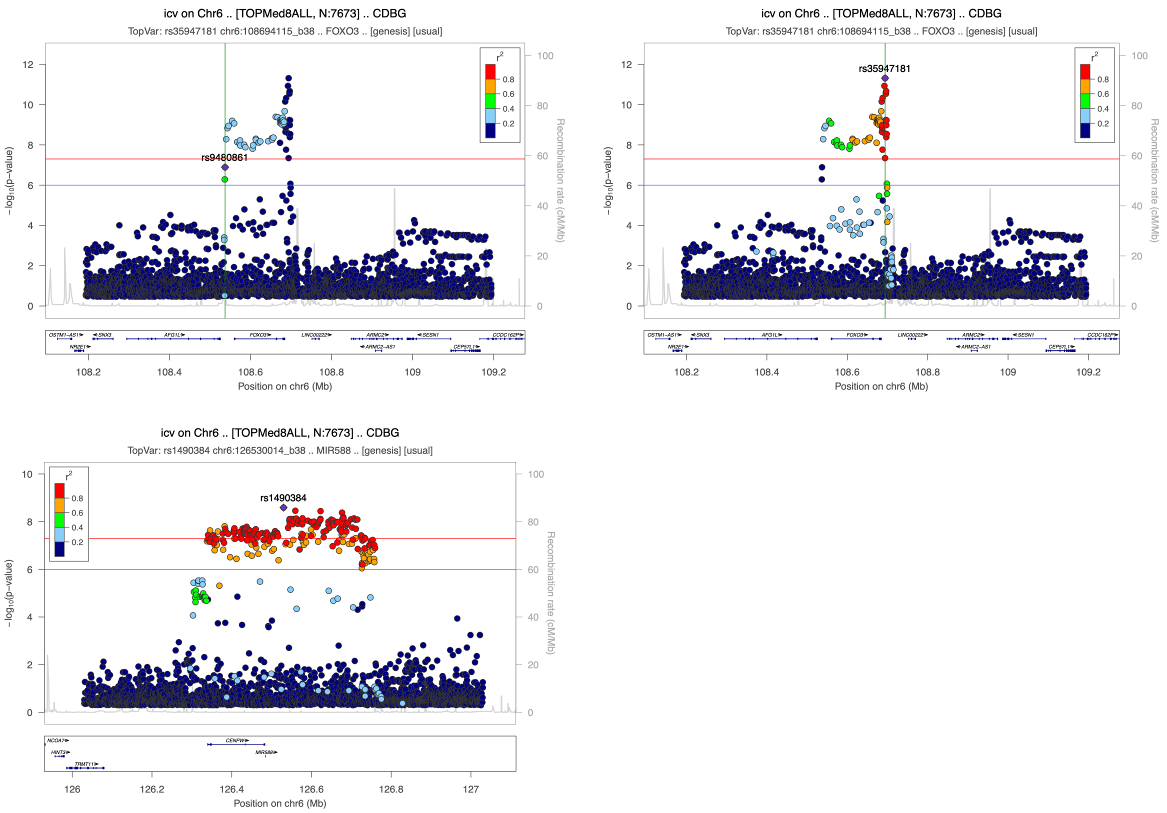


Linkage Disequilibrium (LD) was calculated using the TOPMed reference panel.
Blue and red lines represent significance thresholds of P=$10^{-6}$ and P=$5x10^{-8}$ respectively.

**Supplementary Figure 7**: Regional association plots for HV chr12 (pooled analysis)

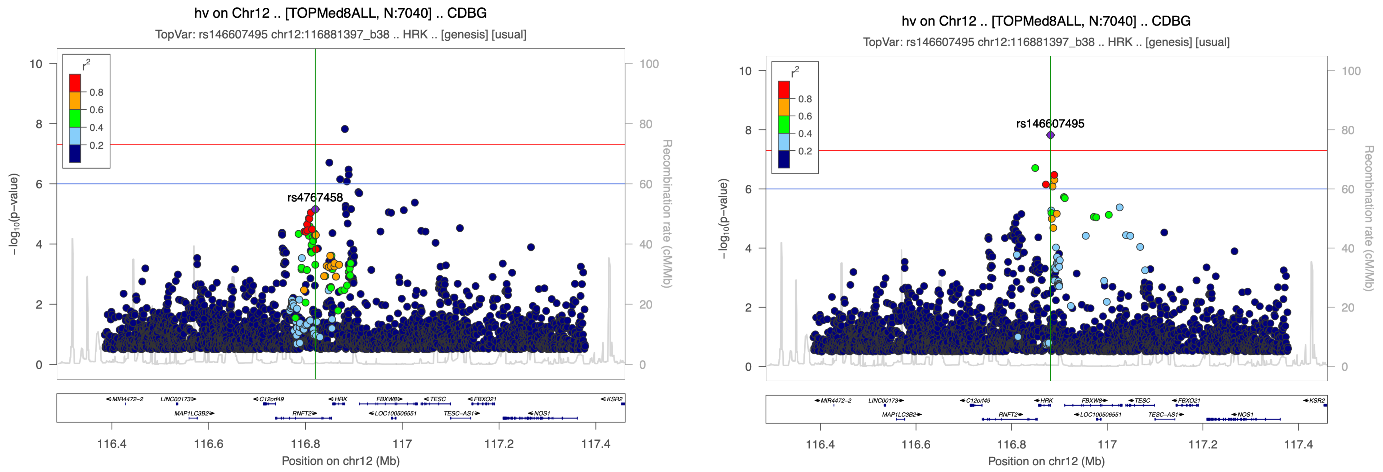


Linkage Disequilibrium (LD) was calculated using the TOPMed reference panel.
Blue and red lines represent significance thresholds of P=$10^{-6}$ and P=$5x10^{-8}$ respectively.

**Supplementary Figure 8**: Regional association plot for LVV chr5 (pooled analysis)

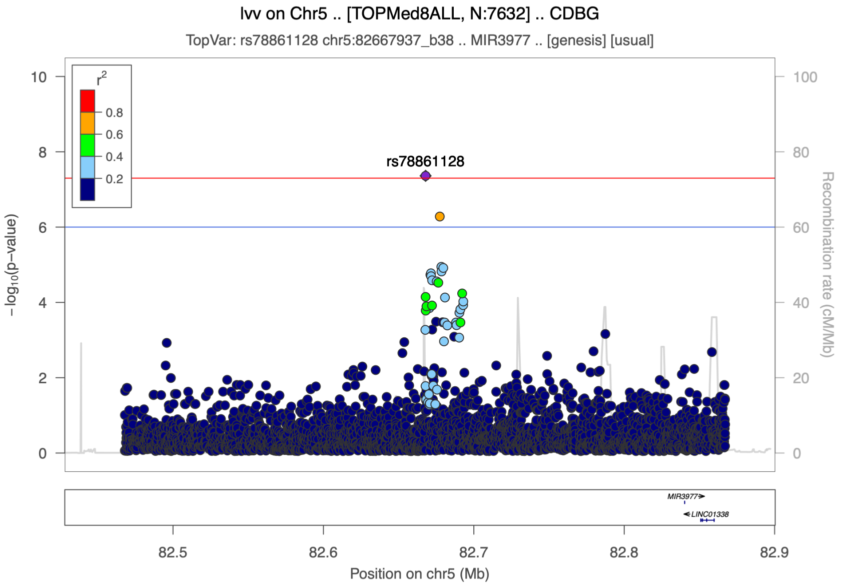


Linkage Disequilibrium (LD) was calculated using the TOPMed reference panel.
Blue and red lines represent significance thresholds of $P=10^{-6}$ and $P=5x10^{-8}$ respectively.

**Supplementary Figure 9**: Regional association plot for LVV chr12 (pooled analysis)

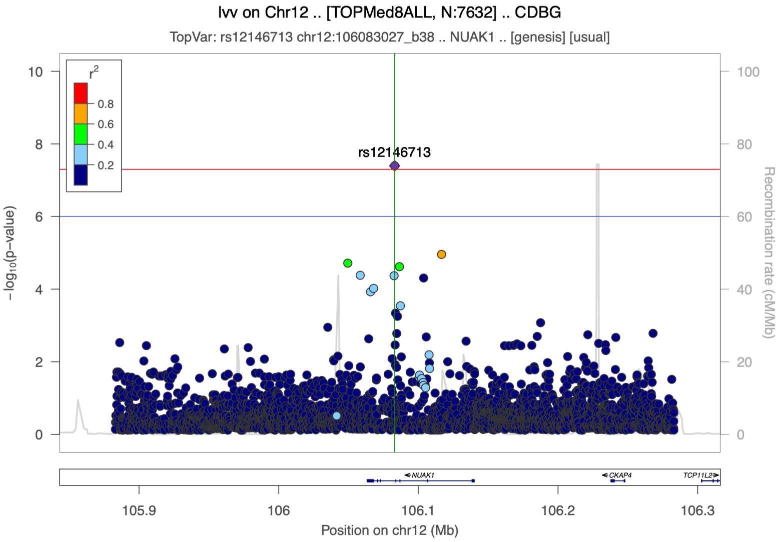


Linkage Disequilibrium (LD) was calculated using the TOPMed reference panel.
Blue and red lines represent significance thresholds of $P=10^{-6}$ and $P=5x10^{-8}$ respectively.

**Supplementary Figure 10**: Regional association plots for HV chr12 from analysis of Black participants

a) Main analysis    b) Conditional analysis on rs1053051

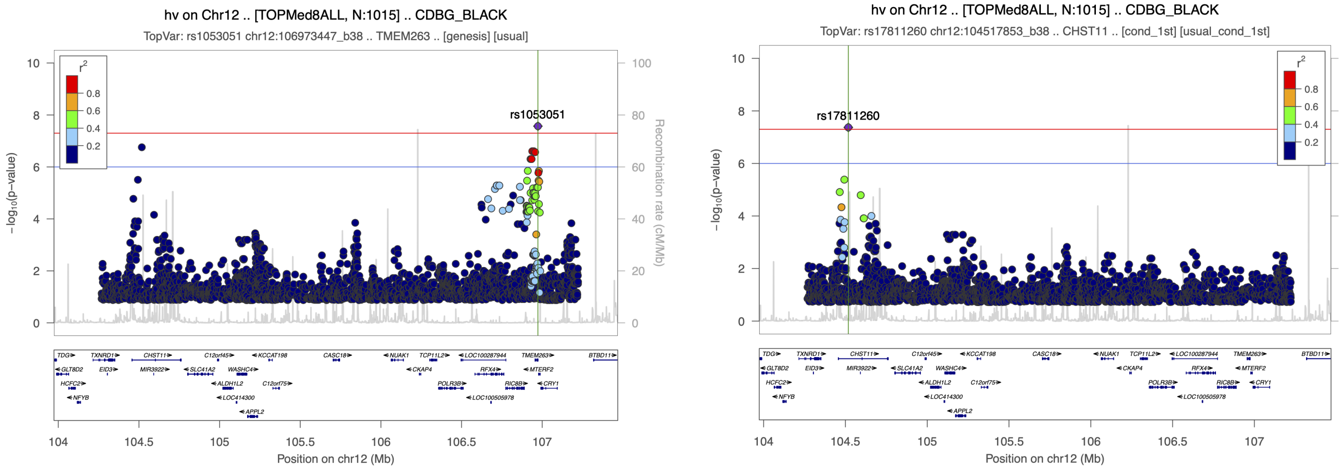


Linkage Disequilibrium (LD) was calculated using the TOPMed reference panel African population. Blue and red lines represent significance thresholds of $P=10^{-6}$ and $P=5x10^{-8}$ respectively.

**Supplementary Figure 11**: Regional association plots for ICV chr18 from analysis of Black participants

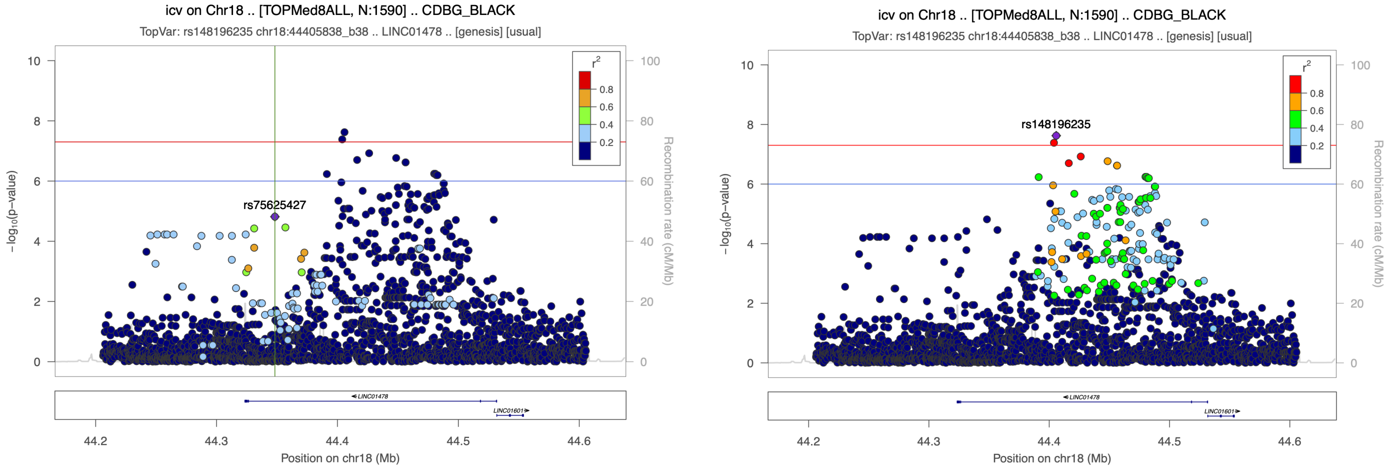


Linkage Disequilibrium (LD) was calculated using the TOPMed reference panel African population. Blue and red lines represent significance thresholds of $P=10^{-6}$ and $P=5x10^{-8}$ respectively.

**Supplementary Figure 12**: Regional association plot for ICV chr20 from analysis of White participants

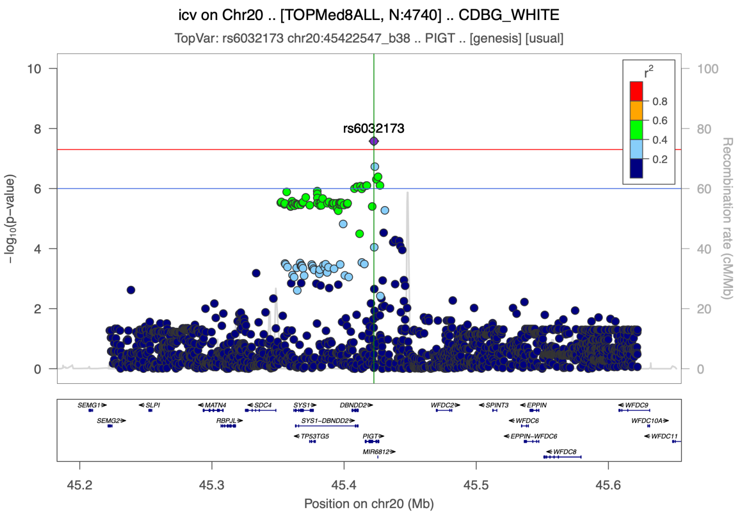


Linkage Disequilibrium (LD) was calculated using the TOPMed reference panel European population. Blue and red lines represent significance thresholds of $P=10^{-6}$ and $P=5x10^{-8}$ respectively.

**Supplementary Figure 13**: Colocalization between Hippocampal volume and *FBXO21* expression in the left ventricle of the heart

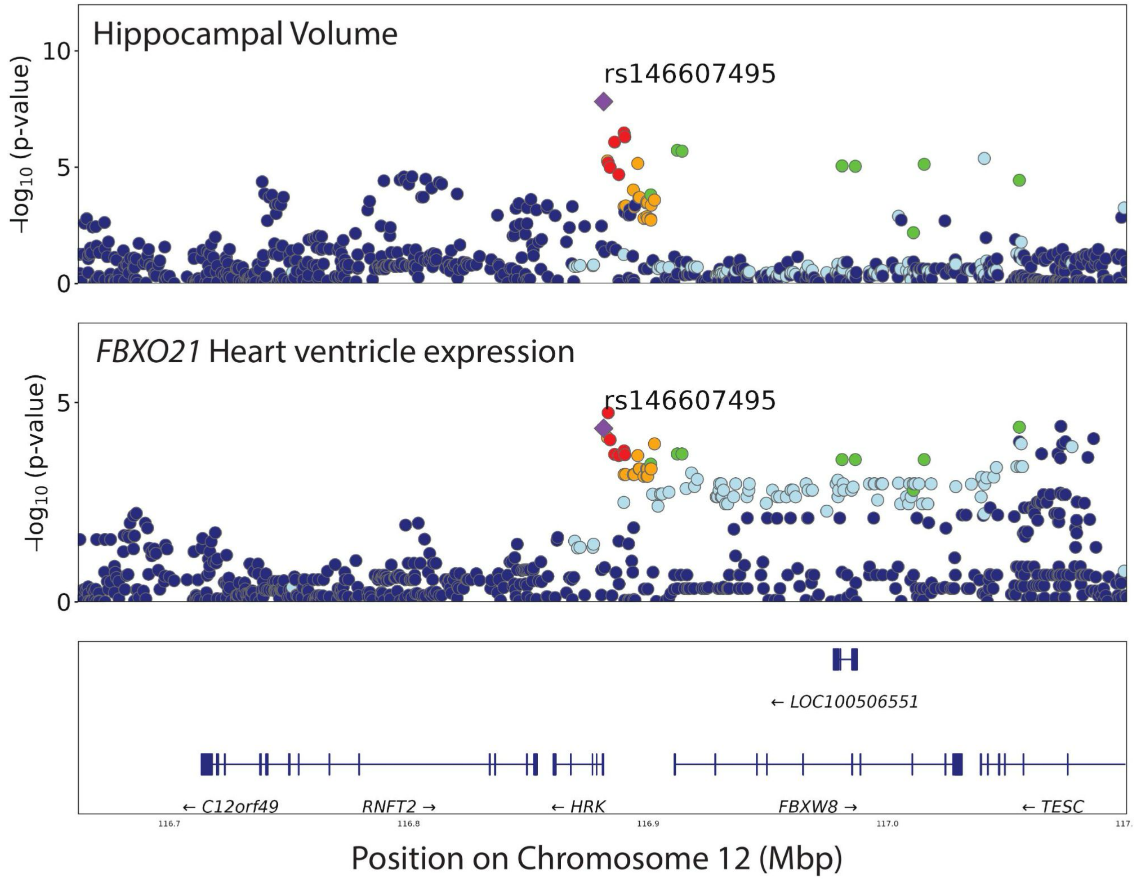


The top two panels show regional plots for hippocampal volume and *FBXO21* expression in the left ventricle of the heart on Chromosome 12 over a 400 kilobasepair span. The bottom panel shows the relative positions of genes in this region. *FBXO21* (not shown) is located 800 Mbp downstream of lead variant rs146607495. *FBXO21* expression summary statistics are from the Genotype Tissue Expression Project (GTEx) v8 release for participants of European ancestry. Colors of dots indicate linkage disequilibrium $r^2$ with lead variant (purple diamond) using 1000 Genomes European (EUR) v3 reference panel: >0.8 (red), 0.6-0.8 (orange), 0.4-0.6 (green), 0.2-0.4 (light blue), <0.2 (dark blue).

**Supplementary Figure 14**: Colocalization between hippocampal volume and antisense to *MTMR9* expression in the nucleus accumbens

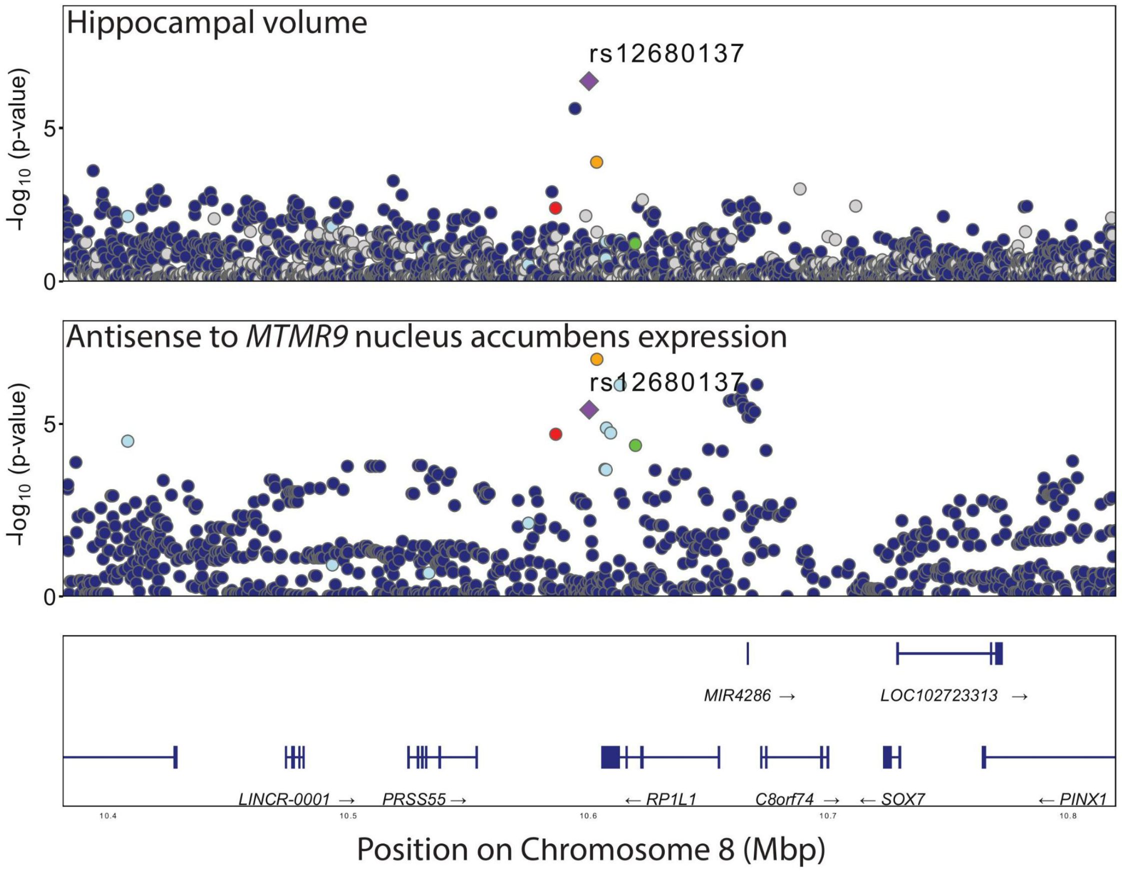


The top two panels show regional plots for hippocampal volume and expression of an RNA gene antisense to *MTMR9* in the nucleus accumbens on Chromosome 8 over a 400 kilobasepair span. The bottom panel shows the relative positions of genes in this region. Antisense to *MTMR9* (not shown) is located 700 Mbp downstream of lead variant rs12680137. Antisense to *MTMR9* expression summary statistics are from the Genotype Tissue Expression Project (GTEx) v8 release for participants of European ancestry. Colors of dots indicate linkage disequilibrium $r^2$ with lead variant (purple diamond) using 1000 Genomes European (EUR) v3 reference panel: >0.8 (red), 0.6-0.8 (orange), 0.4-0.6 (green), 0.2-0.4 (light blue), <0.2 (dark blue).

**Supplementary Figure 15**: Colocalization between lateral ventricular volume and *KIAA1614* expression and methylation in the prefrontal cortex

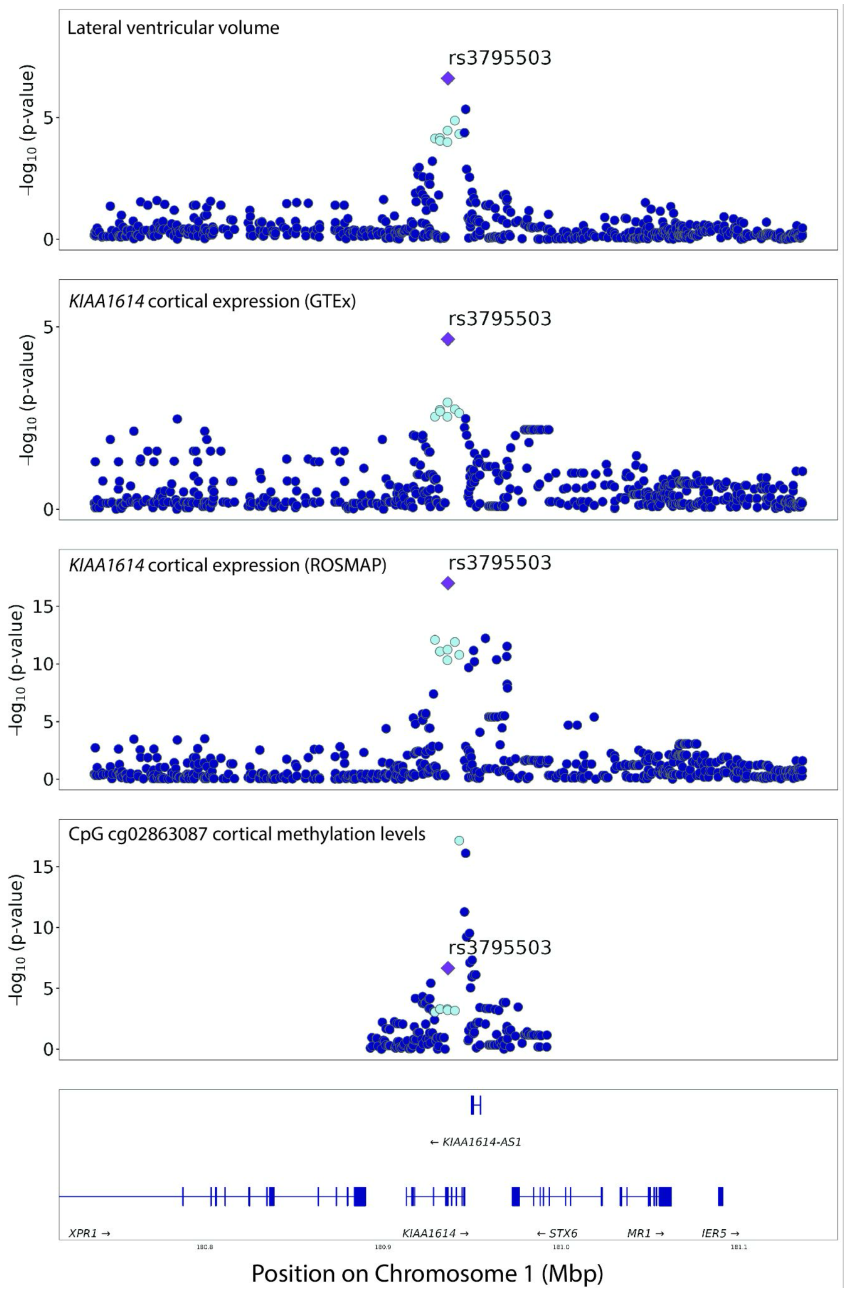


The top four panels show regional plots for lateral ventricular volume, expression of *KIAA1614* in the prefrontal cortex, and methylation of CpG site cg02863087 on Chromosome 1 over a 400 kilobasepair span. The bottom panel shows the relative positions of genes in this region. *KIAA1614* expression summary statistics are from the Genotype Tissue Expression Project (GTEx) v8 release for participants of European ancestry. Colors of dots indicate linkage disequilibrium $r^2$ with lead variant (purple diamond) using 1000 Genomes European (EUR) v3 reference panel: >0.8 (red), 0.6-0.8 (orange), 0.4-0.6 (green), 0.2-0.4 (light blue), <0.2 (dark blue).
